# Disrupted reward processing in Parkinson’s Disease and its relationship with dopamine state and neuropsychiatric syndromes: a systematic review and meta-analysis

**DOI:** 10.1101/2021.10.15.21265008

**Authors:** Harry Costello, Alex J. Berry, Suzanne Reeves, Rimona S. Weil, Eileen M. Joyce, Robert Howard, Jonathan P. Roiser

**Author notes:** Corresponding author: Harry Costello.

## Abstract

**Background:** Neuropsychiatric symptoms are common in Parkinson’s disease (PD) and predict poorer outcomes. Reward processing dysfunction is a candidate mechanism for the development of psychiatric symptoms including depression and impulse control disorders (ICD). We aimed to determine whether reward processing is impaired in PD and its relationship with neuropsychiatric syndromes and dopamine replacement therapy.

**Methods:** The Ovid MEDLINE/PubMed, Embase and PsycInfo databases were searched for articles published up to November 5th, 2020. Studies reporting reward processing task performance by PD patients and healthy controls were included. Summary statistics comparing reward processing between groups were converted to standardized mean difference (SMD) scores and meta-analysed using a random effects model.

**Results:** We identified 55 studies containing 2578 participants (1,638 PD and 940 healthy controls). Studies assessing three subcomponent categories of reward processing tasks were included: Option Valuation (n=12), Reinforcement Learning (n=37) and Reward Response Vigour (n=6). Across all studies, PD patients on medication exhibited a small-to-medium impairment versus healthy controls (SMD=0.34; 95%CI 0.14-0.53), with greater impairments observed off dopaminergic medication in within-subjects designs (SMD=0.43, 95%CI 0.29-0.57). Within-subjects subcomponent analysis revealed impaired processing off medication on Option Valuation (SMD=0.57, 95%CI 0.39-0.75) and Reward Response Vigour (SMD=0.36, 95%CI 0.13-0.59) tasks. However, the opposite applied for Reinforcement Learning, which relative to healthy controls was impaired on-medication (SMD=0.45, 95%CI 0.25-0.65) but not off-medication (SMD=0.28, 95%CI -0.03-0.59). ICD was the only neuropsychiatric syndrome with sufficient studies (n=13) for meta-analysis, but no significant impairment was identified compared to non-ICD patients (SMD=-0.02, 95%CI -0.43-0.39).

**Conclusion:** Reward processing disruption in PD differs according to subcomponent and dopamine medication state and warrants further study as a potential treatment target and mechanism underlying associated neuropsychiatric syndromes.

## Introduction

Parkinson’s disease (PD) is the fastest growing neurological disorder globally,^1^ with estimated annual societal costs comparable to those of dementia.^2^ Traditionally conceptualised as a movement disorder, non-motor symptoms, including disruptions to mood, cognition and motivation are common and have a greater negative impact on health-related quality of life than motor symptoms.^3^ Neuropsychiatric syndromes are common in PD. One-third of patients experience depression,^4^ up to one-half experience apathy^5^ and impulse control disorders (ICDs) associated with dopaminergic medication occur in up to one-quarter.^6^ Currently, there is a lack of understanding of the mechanisms underlying psychiatric symptoms in PD and this represents a barrier to the development of more effective treatments.^7^

Reward processing describes how reinforcement-related perceptions guide goal-directed behaviours^8^. Impaired reward processing is a prominent transdiagnostic feature of several mental health disorders such as depression^8^ and represents a useful framework for understanding symptoms associated with motivation. The National Institute of Mental Health’s Research Domain Criteria (RDoC) identifies reward processing as one of six major domains underpinning human functioning and psychopathology.^9^ Dopamine has a well-established and crucial role in both reward and motivational pathways.^10^ Evidence from dopamine depletion studies has not supported the hypothesis that dopamine mediates hedonic responses (‘liking’), but has revealed a crucial role in motivated behaviours toward desired goals (‘wanting’).^11^

PD is caused by dopaminergic cell death and consequently is a model of striatal and dopamine dysfunction.^12^ The striatum is reciprocally connected with prefrontal areas as well as other parts of the basal ganglia and midbrain forming frontostriatal circuits involved in the initiation and control of motor, cognitive and emotional behaviour. These pathways also constitute part of the brain’s reward circuit, responsible for modulating reward-related behaviour and learning.^13^ Psychiatric syndromes in PD are thought to reflect dysfunction of non-motor frontostriatal circuitry; for example ICDs, are believed to develop through aberrant reward processing, due to an interaction between the disrupted reward processing circuitry underlying PD and dopamine agonist treatment.^14^

Over the past two decades, studies of reward processing in PD have typically used behavioural tasks assessing three subcomponent processes of reward processing:^8^ (1) Option Valuation, the process by which individuals evaluate reward-related options when given explicit information about those options (e.g., reward, cost, and probability); (2) Reward Response Vigour, which reflects the speed or strength with which an individual executes an action to obtain a reward; (3) Reinforcement Learning, which describes the process by which an individual uses feedback to change their future behaviour. To date, there has been one meta-analysis of Iowa Gambling Task performance in PD, which reported significantly impaired reward learning.^15^ However, the degree and pattern of impairments on other reward processing tasks in PD and any relationship with dopaminergic state and psychiatric symptoms remain unclear.

Here we report the first systematic review and meta-analysis of reward processing behaviour in PD and its relationship with dopamine replacement therapy and associated neuropsychiatric syndromes. Our aims were: (1) To clarify the nature and extent of differences across reward processing subcomponents between PD and healthy groups; (2) To test the role of dopamine state (on or off medication) in reward processing in PD; (3) To investigate any differences in reward processing in PD patients with and without neuropsychiatric syndromes.

## Method

### Systematic review

The Ovid MEDLINE/PubMed, Embase, and PsycInfo databases were searched for articles published between January 1st, 1946, and November 5th, 2020 inclusive, with titles or abstracts containing the terms: Parkins* and (reward* or motivat* or incentiv* or effort* or deci*) and (psychiatric or neuropsychiatric or depress* or psychosis or delus* or impuls* or mood or anxiety or apathy or anhedonia or hallucin*). Inclusion criteria were as follows: (1) case-control design; (2) included a group with Parkinson’s disease without dementia or deep brain stimulation; (3) participants were at least 18 years old; (4) participants performed a reward-processing task; (5) task rewards were explicit, i.e. money, points, water or food (we did not include studies that used outcomes that could be considered purely informational or social feedback, e.g. happy/sad faces or variants of correct/incorrect, to ensure specificity); (6) studies reported data on a behavioural measure of reward processing that could be converted to a case-control standardized mean difference (SMD) score. If this was not reported, data were requested from the authors. Articles were independently assessed by H.C and A.B, using a rating tool based on the Newcastle-Ottawa scale^16^ for assessing the quality of nonrandomized studies (Supplement). Conflicts in quality assessment rating were resolved through in-person discussion.

### Meta-analysis

Behavioural measures from each study were categorised as measuring Option Valuation, Reward Response Vigour, or Reinforcement Learning, and converted to an SMD score and an associated standard error (see Supplement for equations).^17^

Within the Option Valuation and Reward Response Vigour subcategories, a positive SMD represents a greater or faster response to reward by the control than the PD group, respectively. A positive SMD within the Reinforcement Learning subcategory represents faster use of feedback to maximize reward by the control group than the PD group.

Meta-analysis was conducted if four or more studies were present within a reward processing subcategory for PD patients compared with healthy controls, PD with and without a psychiatric symptom, or PD on-compared with off-medication (within-subjects designs only).

Meta-analysis was performed using the R statistical programming language and the packages metafor and metaviz, using random effects models. Heterogeneity was analysed using the approximate proportion of total variability (I^2^).

Funnel plot asymmetry was assessed using visual inspection of a contour-enhanced funnel plot and the Egger test.

## Results

We initially identified 2,122 studies, excluded 1,898 of these by title/abstract and retrieved the remaining 224 full papers (Figure 1). Data from 55 studies containing 2,578 participants (1,638 PD, 940 healthy controls) were analysed (see PRISMA diagram in Figure 1), two studies could not be used in the quantitative analysis due to a lack of reported summary statistic figures. The median number of patients per study was 24 (IQR 16), median participant age was 63.3 years (IQR 7.5) and median duration of PD was 7.0 years (IQR 4.5).

**Figure 1.**
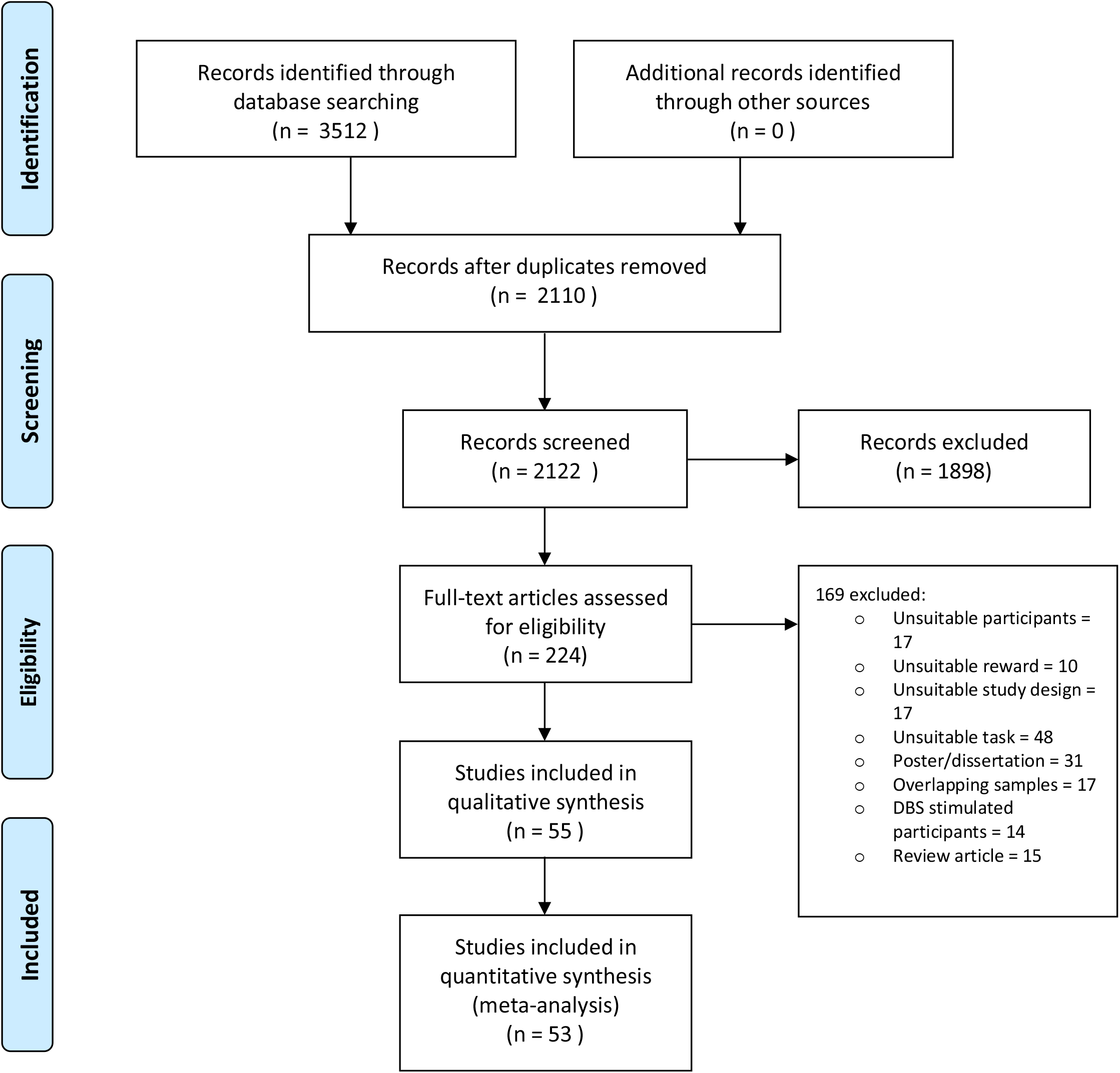
PRISMA flow diagram of study selection and inclusion.

Meta-analysis across all reward processing subcomponent categories identified a small-to-medium reward processing impairment in PD patients both on- (SMD=0.34; 95%CI 0.14-0.53) and off-medication (SMD=0.40; 95%CI 0.19-0.62), compared to healthy controls (Figure 2a,b). Within-subjects comparison of reward processing between on- and off-medication states was possible in 14 studies, revealing relatively impaired reward processing off-medication, with a medium effect size (SMD=0.43, 95%CI 0.29-0.57; Figure 2c).

**Figure 2.**
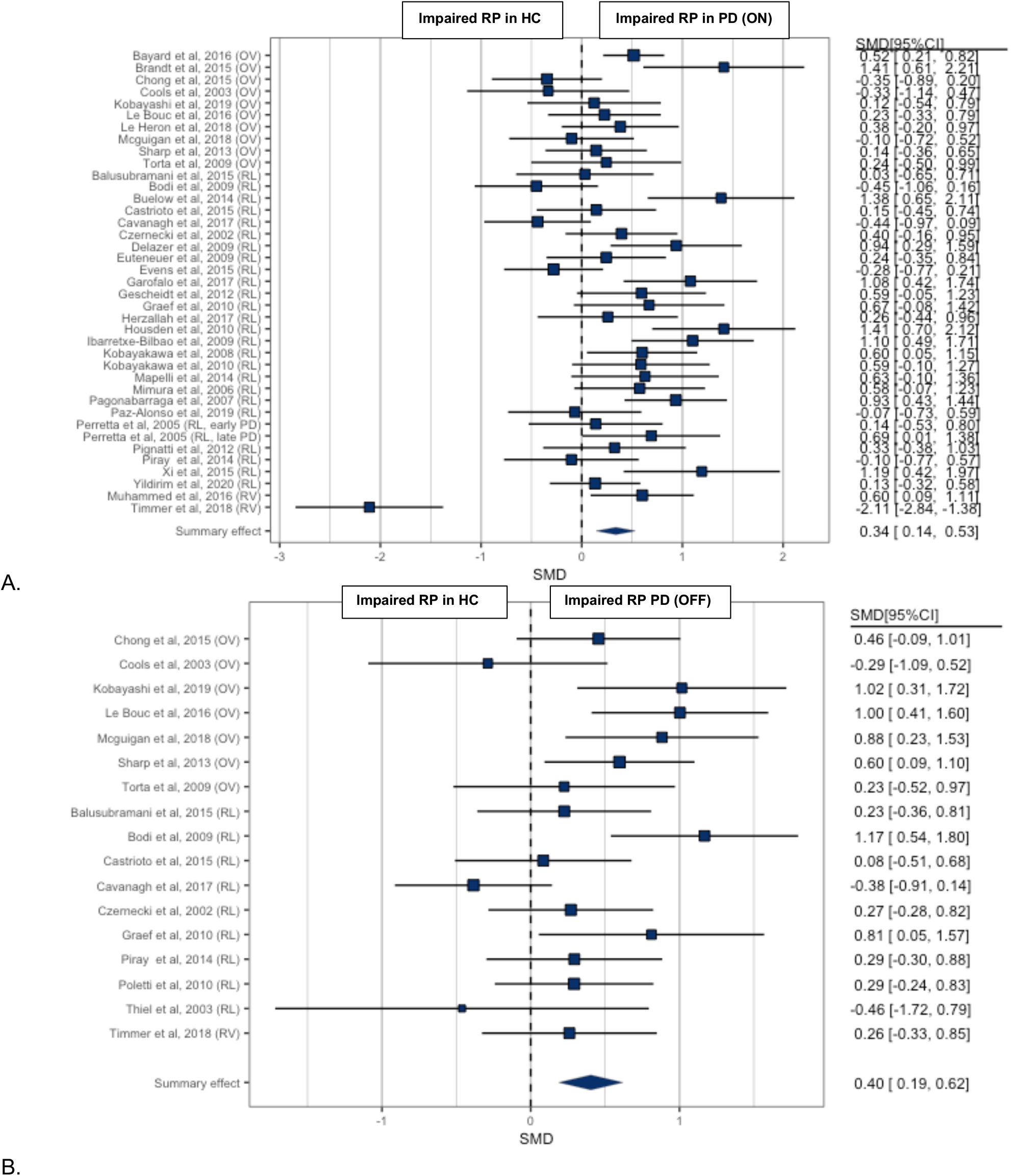
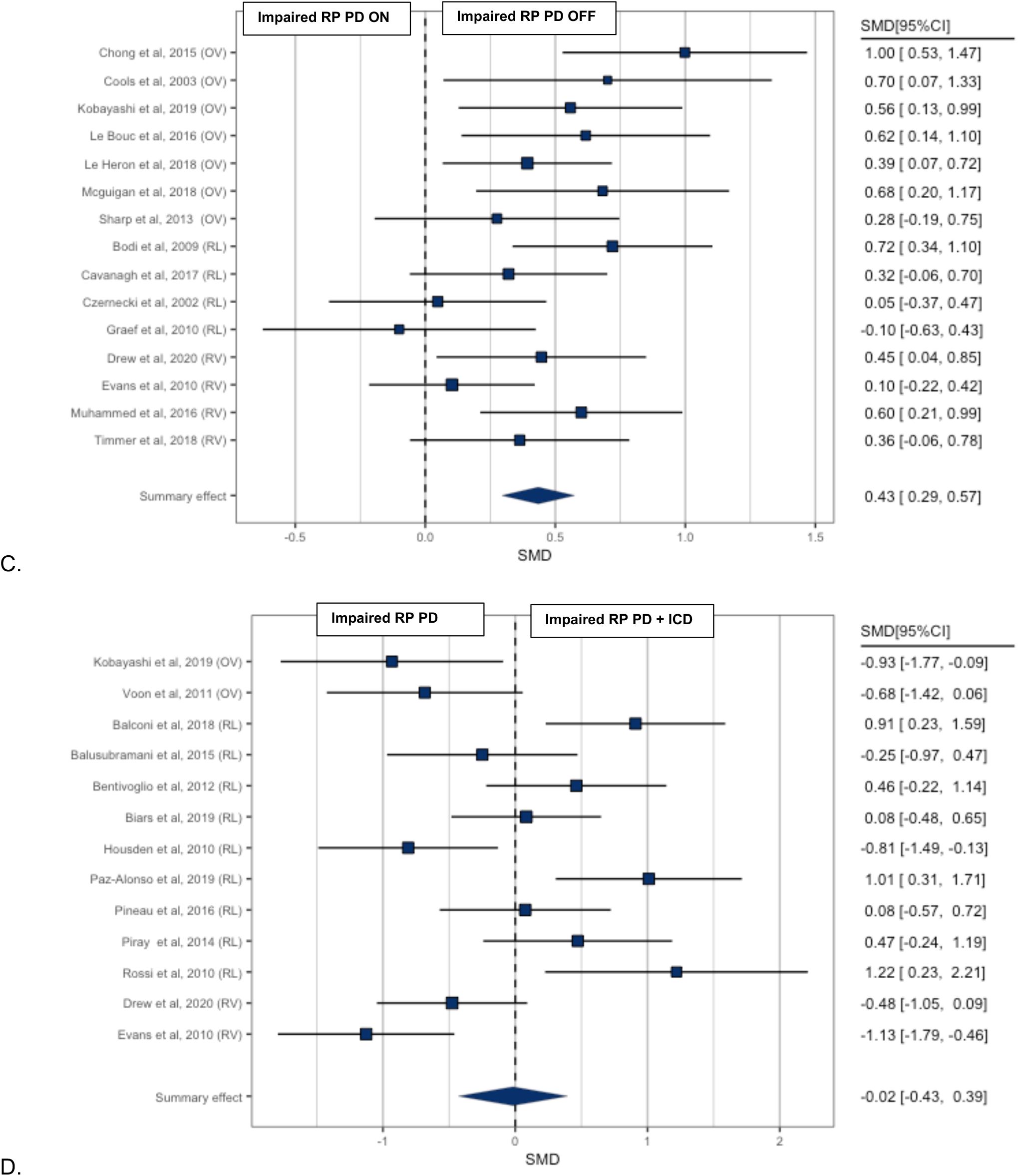
Forest plot of reward processing (RP) in (A) PD ON versus health controls (HC) (B) PD ON versus health (C) the ON versus OFF dopamine state (D) PD with and without Impulse Control Disorder (ICD)

ICD was the most studied and only neuropsychiatric syndrome with sufficient studies (n=13) for meta-analysis. No significant impairment (see Figure 2d) was identified in reward processing in PD patients with ICD compared to non-ICD patients (SMD=-0.02, 95%CI -0.43-0.39).

Overall interstudy heterogeneity was substantial (I^2^=57.48%), and the median power of included studies and *R*-index was low (Supplement Figure 1, median power=36%; R index=28%). Analysis of funnel plot asymmetry using Egger’s regression line did not meet statistical significance (p=0.32) and was likely a consequence of high heterogeneity and small sample size of included studies.

Quality assessment and risk of bias analysis using a modified Newcastle-Ottawa scale (see Supplement Table 7) found the majority of included studies used a validated assessment tool for diagnosis of PD (65.5%), and accounted for PD severity (94.5%) and medication status (90.9%).

However, almost half of included studies gave no description of how healthy controls were selected (42.2%) or clearly defined controls as having no past psychopathology (42.2%).

### Option Valuation

We identified 12 studies containing 347 PD patients and 278 healthy participants that used Option Valuation tasks (Supplement Table 1). The mean age of participants was 62.9 (±4.6) years, and mean duration of illness was 7.5 (±2.8) years. Effort-based decision-making tasks (three studies) and the game of dice task (3 studies) were most commonly used. Four studies reported psychiatric medication use in participants, three of which included participants taking antidepressant medications.

Meta-analysis of studies comparing Option Valuation in PD patients compared with healthy controls showed lower reward weighting in PD, which was moderated by dopamine medication (Figure 3a&b). Patients on-medication did not differ significantly from healthy controls (SMD=0.22, 95%CI -0.04– 0.49), but off-medication there was a medium-to-large impairment (SMD=0.60, 95%CI 0.30-0.89). Within-subjects comparison confirmed lower relatively reward weighting off-medication, with a medium-to-large effect (SMD=0.57, 95%CI 0.39-0.75; Figure 3c).

**Figure 3.**
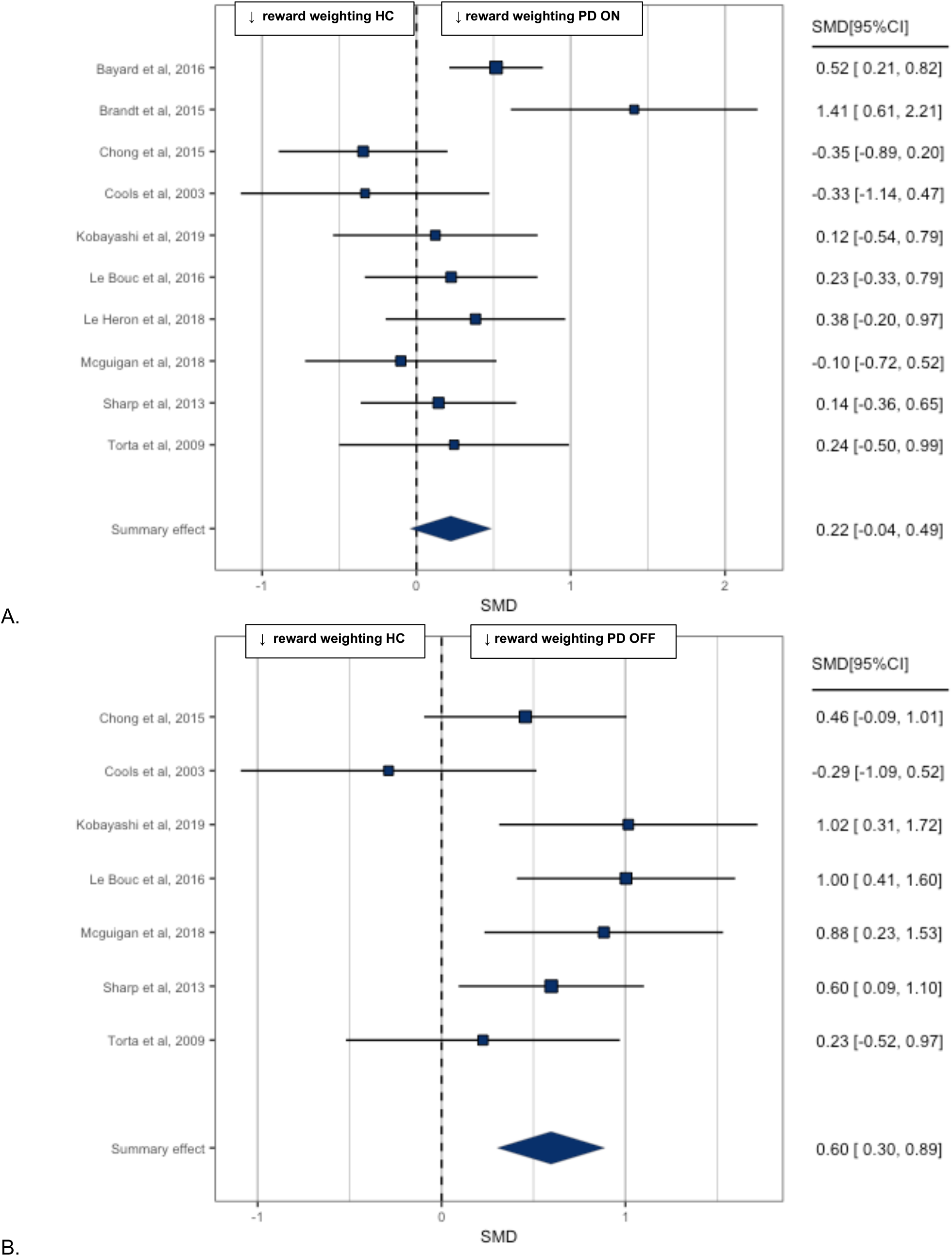
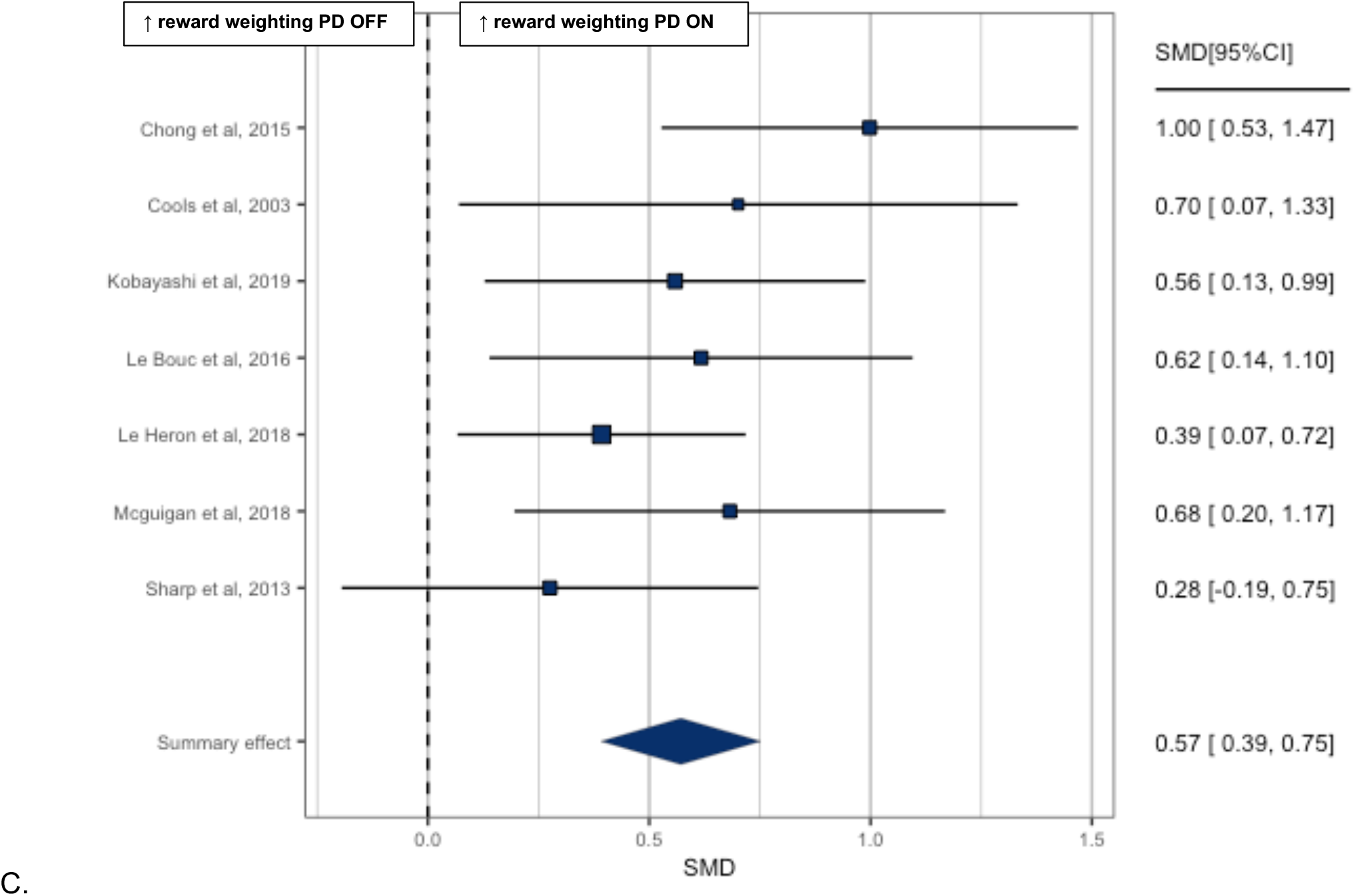
Forest plot of Option Valuation in: (A) PD ON versus health controls. (B) PD OFF versus health controls. (C) The ON versus OFF dopamine state.

Four studies compared Option Valuation in PD patients with and without neuropsychiatric syndromes. Three of these studies^18–20^ compared option valuation in PD patients with and without ICD, with mixed findings. One study^19^ using an economic choice task reported lower reward weighting in ICD, while the other two studies^18,20^ using gambling tasks found no difference^18^ and increased reward weighting,^20^ respectively.

One study^21^ investigating the effect of apathy on Option Valuation reported lower acceptance of offers of reward obtained through physical exertion. This pattern of impairment in apathy was found to be dissociable from the effects of dopamine. Apathy was characterised by rejection of predominantly low reward offers, while dopamine state mediated response to high effort, high reward offers.

In summary, Option Valuation impairment in PD is dopamine dependent, with lower reward weighting off dopaminergic medication. Too few studies have investigated Option Valuation in PD patients with neuropsychiatric syndromes to draw meaningful conclusions.

### Reinforcement Learning

We identified 37 studies containing 1,059 PD patients and 593 healthy controls that used Reinforcement Learning tasks (Supplement Table 2). The majority of studies (20/37) used the Iowa Gambling Task. Ten studies reported psychiatric medication use, of which three included participants taking antidepressant medication.

Reinforcement Learning was slowed in PD patients on-medication versus healthy controls (Figure 4a&b) with a medium effect size (SMD=0.45, 95%CI 0.25-0.65). Interestingly, there was no significant group difference off-medication (SMD=0.28, 95%CI -0.03-0.59). Comparison of Reinforcement Learning comparing on- and off-medication within-subjects (Figure 4c) was possible in four studies, which did not detect a significant effect (SMD=0.27, 95%CI -0.08-0.62); however, we note that this analysis is likely underpowered due to the small number of included studies.

**Figure 4.**
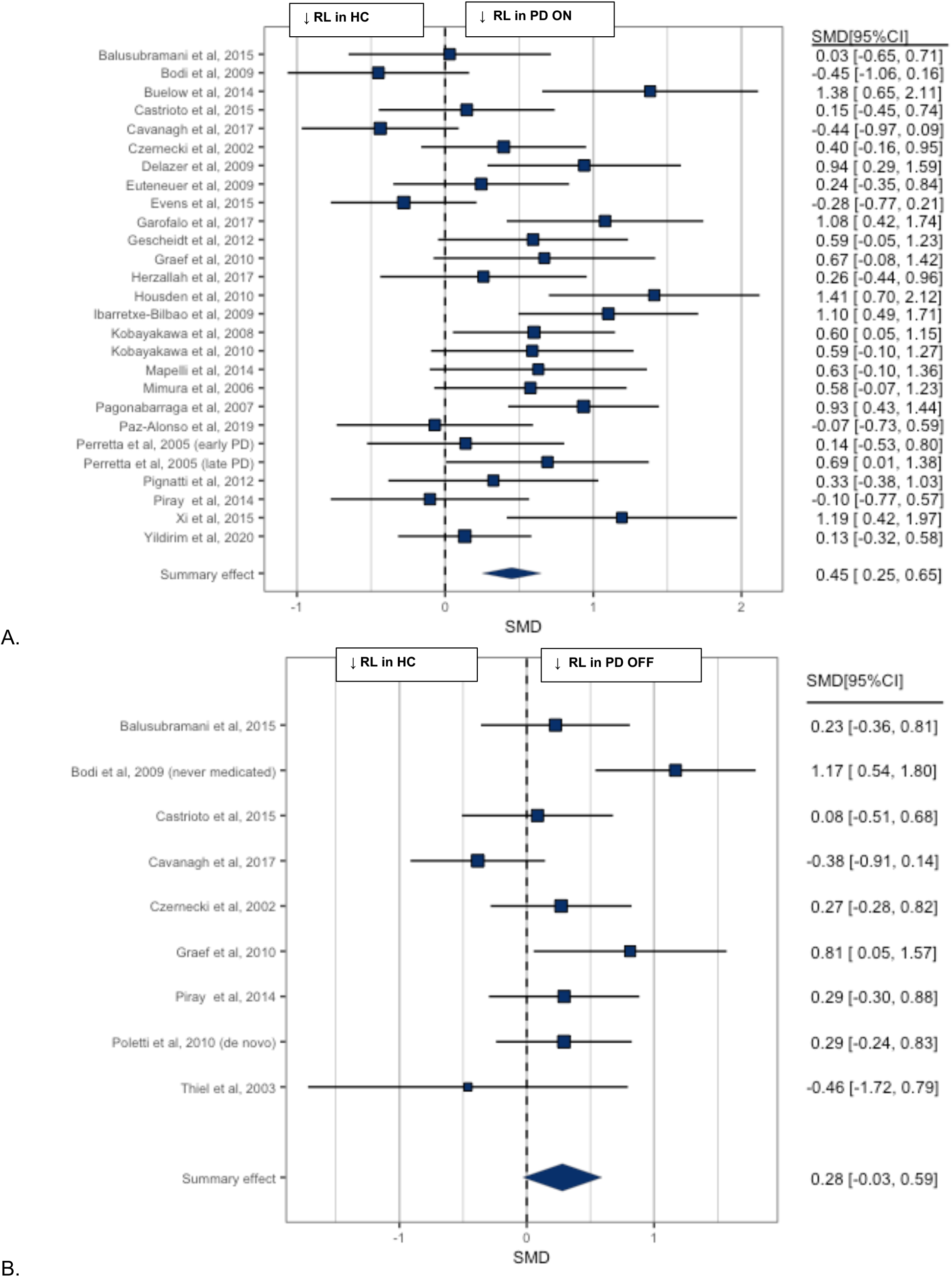
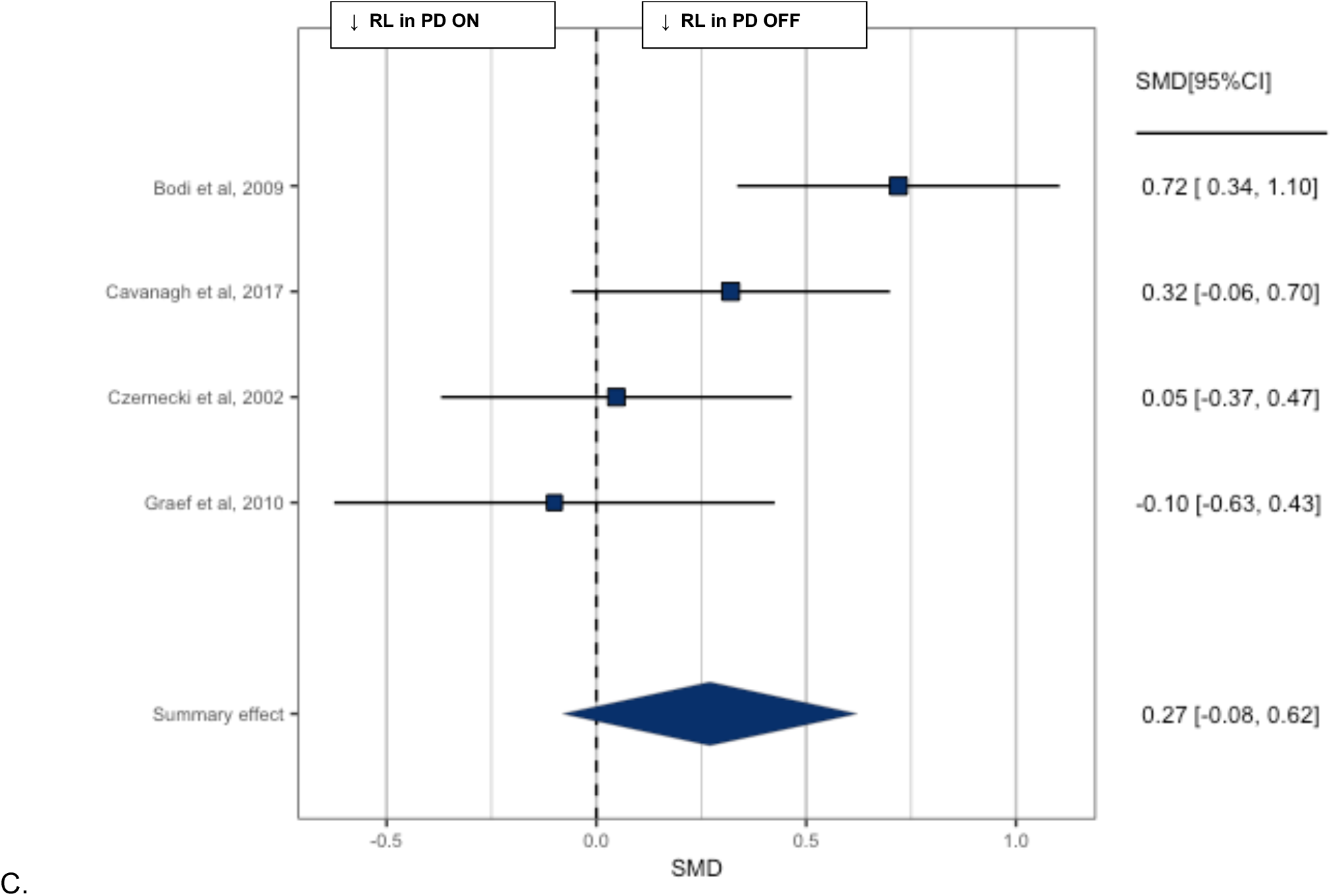
Forest plot of Reinforcement Learning (RL) in: (A) PD ON versus health controls. (B) PD OFF versus health controls. (C) The ON versus OFF dopamine state.

Sixteen studies investigated reinforcement learning in PD patients with and without neuropsychiatric symptoms (Supplement Table 2), with the majority (11/16) examining ICD. Meta-analysis of nine studies (Supplement Figure 2) found no significant difference between ICD and non-ICD PD patients (SMD=0.32, 95%CI -0.09-0.73).

Two studies^22,23^ examined Reinforcement Learning in PD patients with major depressive disorder. Both ^22,23^ reported impaired Reinforcement Learning in depressed compared to non-depressed PD patients. One^23^ also compared Reinforcement Learning in depressed PD patients with depressed participants without PD. A similar pattern of impairment in learning from positive feedback was identified in the two groups, suggesting that Reinforcement Learning impairment may not be specific to depression in PD.^9^

Two studies^24,25^ examined the role of apathy in reward learning. Both used the Iowa Gambling Task but reported conflicting findings: one found significant impairment^25^ but the other reported better reinforcement learning in PD patients with apathy,^24^ compared to those without.

In summary, and in stark contrast to studies of Option Valuation, Reinforcement Learning is particularly impaired in PD in the on-medication state. There was no significant impairment in Reinforcement Learning in PD patients with ICD compared with those without ICD. Too few studies have investigated Reinforcement Learning in PD patients with other neuropsychiatric syndromes to draw meaningful conclusions.

### Reward Response Vigour

We identified seven studies containing 232 PD patients and 69 healthy controls that investigated Reward Response Vigour in PD (Supplement Table 3). Insufficient studies were identified to allow meta-analysis of Reward Response Vigour in PD compared with healthy controls. Of the three studies^26–28^ that reported reward-related speeding in PD and healthy controls, results were mixed, with studies reporting lower,^26^ greater^27^ and no difference^28^ in PD patients compared to healthy volunteers.

Meta-analysis of the effect of dopamine state on Reward Response Vigour in four studies (Figure 5) identified a small-to-medium increase in Reward Response Vigour on-medication (SMD=0.36, 95%CI 0.13-0.59).

**Figure 5.**
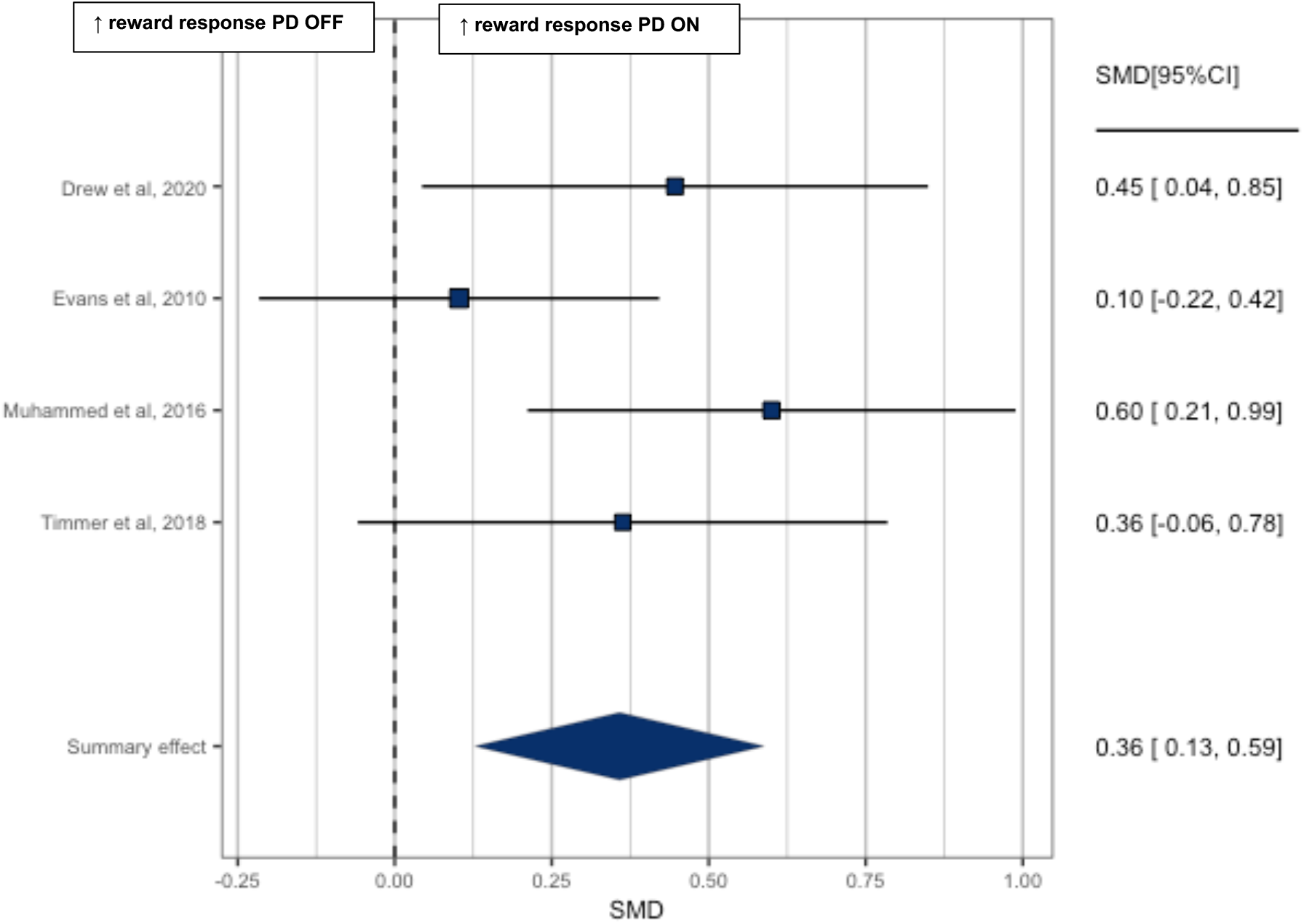
Forest plot of Reward Response Vigour in Parkinson’s disease ON versus OFF dopamine state.

Six studies investigated Reward Response Vigour in PD patients with and without neuropsychiatric syndromes (Supplement Table 3). Two studies^27,29^ examined apathy, one using a rewarded saccadic eye movement task,^27^ the other a rewarded spatial search task;^29^ both reported no significant group differences. Similarly, no significant difference in Reward Response Vigour was found in two studies comparing ICD and non-ICD patients^30,31^, and two investigating depression in PD^28^.

In summary, relatively few studies have investigated Reward Response Vigour in PD, and findings are mixed. Reward Response Vigour in PD was reduced in the off-compared to the on-medication state. Too few studies have investigated Reward Response Vigour in PD patients with neuropsychiatric syndromes to draw meaningful conclusions.

## Discussion

This is the first systematic review and meta-analysis of reward processing in PD, associated neuropsychiatric syndromes and the influence of dopaminergic medication. Across all 55 studies, including different subcomponents of reward processing, we found PD patients to have small-to-medium reward processing impairments relative to healthy participant groups. The degree of impairment in reward processing is similar to that reported in major depressive disorder, a condition where dysfunctional reward processing is a leading aetiological candidate mechanism for “interest-activity” symptoms, such as anhedonia.^8^ We also identified potentially important differences between reward processing subcomponent categories and the effect of dopamine state.

The Option Valuation subcategory exhibited the largest impairment in PD which was dopamine dependent, with markedly reduced reward weighting in PD patients off dopaminergic medication. This finding is supported by animal^11^ and human experimental studies^32^ which show impaired valuation following dopamine depletion. Dopamine antagonists such as antipsychotic drugs also reduce preference for high-effort/high-reward options,^11^ suggesting that dopamine transmission is crucial in cost-benefit decision making. Dopaminergic pathways in the brain reward circuit including the anterior cingulate cortex and basal ganglia are believed to be central in choosing and executing effortful action.^5^ Option Valuation is a component of effort-based decision making and represents a framework for understanding apathy and anhedonia, both common motivational disorders in PD and depression.^5^ However, no study to date has investigated Option Valuation in depression in PD, and the only study^21^ to examine apathy found dissociable effects of dopamine and apathy on decision making, indicating impairment may not only be secondary to dopamine depletion.

In direct contrast to the pattern identified in the Option Valuation subcategory, Reinforcement Learning was moderately impaired in PD when patients were *on* dopamine medication, with no significant difference detected when off medication. This is surprising given decades of evidence that dopaminergic pathways from the midbrain are crucial for reward learning.^33^ However, recent studies applying cell-type specific monitoring and manipulation of distinct neuronal populations in the striatum have suggested that heterogenous signals in dopaminergic neurons support specific types of learning.^34^ For example, differentially regulated mechanisms of dopamine release in the basal ganglia underlie distinct functions.^35^ Reward learning is believed to be facilitated by dopamine cell spiking encoding reward prediction errors, whereas gradual increase in dopamine release mirrors reward expectation^35^. Reinforcement learning is therefore believed to be dependent on phasic rather than tonic dopamine signalling. Wave-like spatiotemporal dopamine dynamics in the dorsal striatum have also been implicated in encoding reward prediction errors to facilitate learning.^36^ It remains unclear what effect exogenous dopamine in PD has on the dynamics of striatal dopamine signalling. Studies of associative learning in healthy subjects have found that dopamine agonists can impair learning by inhibiting phasic dopamine signalling.^37^ Therefore, one possible interpretation is that dopamine medication may remediate control of reward expectation and motivation within the striatum, but impair the broadcast burst signals required to promote learning.^35^ However, this requires testing in future studies.

Distinct types of reinforcement learning model utilised during task performance may also play a crucial role.^38^ ‘Model-free’ learning describes learning through direct experience rather than through constructing an internal model of the environment in order to develop a complex map of cues and actions which lead to reward.^38^ Most studies included in our review used model free reinforcement learning tasks. Evidence suggests that these two types of reinforcement learning processes are mechanistically distinct, and differentially dependent on dopamine reward prediction errors.^38^

The Reward Response Vigour subcategory showed a significant small-to-moderate impairment in the off -compared to the on-medication state in PD patients. However, relatively few studies were identified and reaction times may be vulnerable to attentional confounds. Though several studies reported reaction times during tasks, reward related speeding (i.e. the difference between rewarded and non-reward conditions) was infrequently measured, without which slower reaction times would likely only reflect bradykinesia associated with PD.

Despite PD being a model for dopamine dysfunction, current treatments of common neuropsychiatric syndromes in PD such as depression do not differ from depression in patients with other long-term conditions^39^ and have limited efficacy.^40^ Symptoms of anxiety and depression in PD are mostly present in the off-dopamine state,^41^ suggesting depression in PD may be related to dopaminergic deficit and have a specific aetiology. Our findings suggest PD is characterised by a specific pattern of impairment in reward processing which is dopamine dependent and potentially could be a causal mechanism underlying neuropsychiatric symptoms such as depression. However, other than ICD, which we found was not significantly associated with reward processing impairment, few studies have investigated reward processing in PD-associated neuropsychiatric syndromes. Further understanding of how impairment in reward processing is associated with specific neuropsychiatric manifestations of PD is needed to understand the underlying mechanisms of these disabling syndromes and develop more targeted and effective treatments.

### Limitations

We categorised reward processing into three subcomponent categories, however there are several ways to measure function in each category which grouped diverse processes. For example, the Option Valuation subcategory included studies measuring risk taking and decisions to exert effort, resulting in meta-analysis of heterogeneous measures. A minority of studies reported psychiatric medication use in participants. Evidence suggests antidepressant medication may partly exert its effect via modulating reward processing^42^ which could have confounded results. Though we measured and compared the effect of dopamine medication state on task performance, the medication regime and proportion of patients on dopamine agonist treatment as opposed to levodopa was reported in less than half of included studies (22/55). Different PD medications are disproportionately associated with dopamine related psychiatric conditions such as ICD,^6^ and distinct regimes could potentially impact reward processing variably. The majority of studies investigating reward processing in PD-associated neuropsychiatric syndromes used PD patients without the syndrome as a control group. Only one study^23^ investigating depression in PD used a control group of patients with depression without PD. In order to establish whether patterns of reward processing impairments are specific to PD-associated neuropsychiatric syndromes and not a common feature of psychiatric symptoms more generally, further studies of this type are needed. Finally, our systematic review and meta-analysis examined the findings of case-control studies which are unable to inform us of the causal relationship between reward processing impairment, PD and its associated neuropsychiatric syndromes. Longitudinal studies are needed to answer these questions and understand how reward processing changes develop as PD advances. Our analyses of the impact of dopamine medication were derived from studies conducted using within-subjects experimental comparisons, and therefore we can be more confident of a causal role. However, the effects of being off-medication in a patient who usually takes dopamine-boosting drugs, including heightened anxiety and physical discomfort, could plausibly affect task performance. A minority of studies (22/55) measured motor symptom severity in both on and off states, and only four studies measured differences in anxiety symptoms in both states.

## Conclusions

PD is associated with a small-to-medium level of reward processing impairment overall, with variable degrees of impairment across subcomponent reward processing categories. Reward processing is dependent on dopamine state with greater impairment in Option Valuation and Reward Response Vigour when patients are off dopaminergic medication, but surprisingly faster Reinforcement Learning. Other than Reinforcement Learning in ICD, few studies have investigated the relationship between reward processing and PD associated neuropsychiatric syndromes. Further research, including longitudinal studies are needed to conclude whether specific patterns of impairment in reward processing have a causal relationship with neuropsychiatric syndromes in PD.

## Data Availability

All data produced in the present work are contained in the manuscript.

## Contributorship

Study concept and design: HC, JR and RH. Acquisition of data: HC, AJB and JR. Analysis and interpretation of data: HC and JR. Drafting of manuscript: HC All authors critically revised successive drafts of the paper and approved the final version.

## Ethical approval

This study is secondary research that synthesised the results of original papers; as such, it is exempt from ethical approval.

