## Supplementary material for "Disrupted reward processing in Parkinson’s Disease and its relationship with dopamine state and neuropsychiatric syndromes: a systematic review and meta-analysis"

Supplement figure 1. Contour-Enhanced Funnel Plot of all studies Parkinson's versus healthy controls

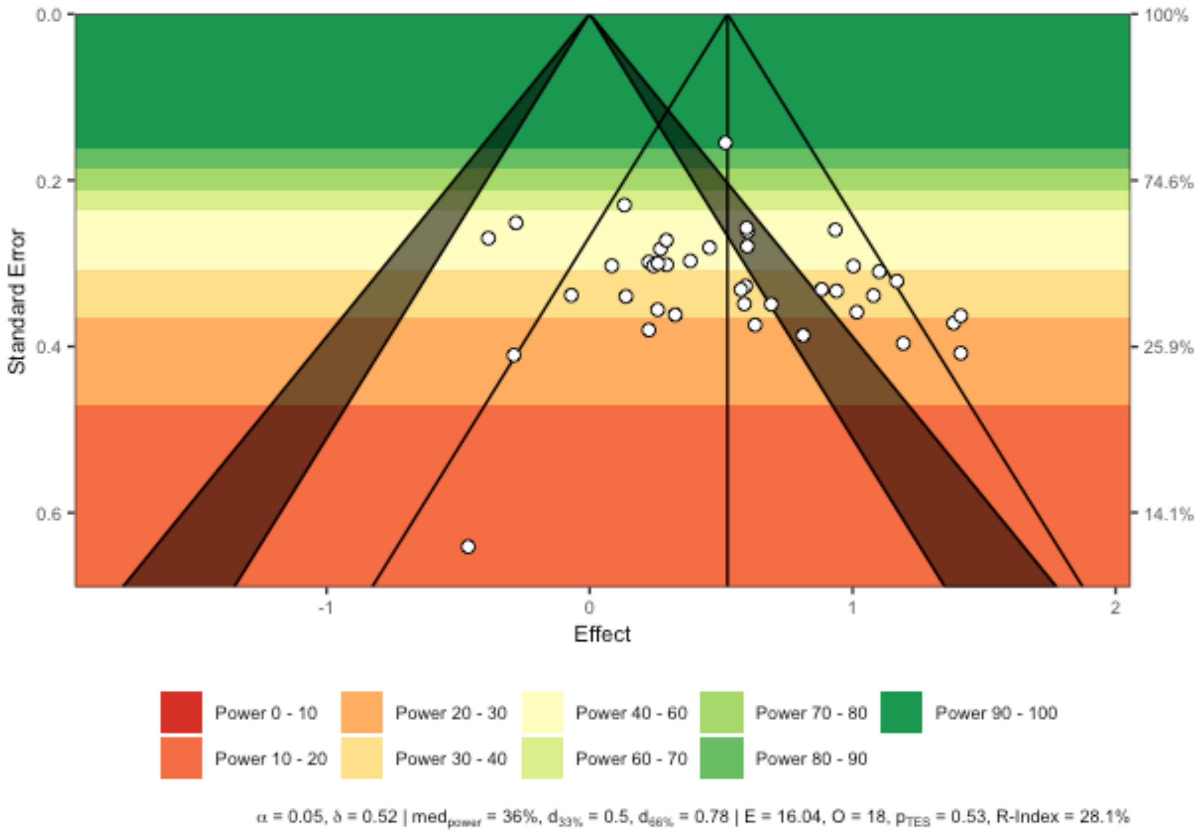

Supplement figure 2. Forest plot of reinforcement learning (RL) in Parkinson's patients with and without Impulse Control Disorder (ICD)

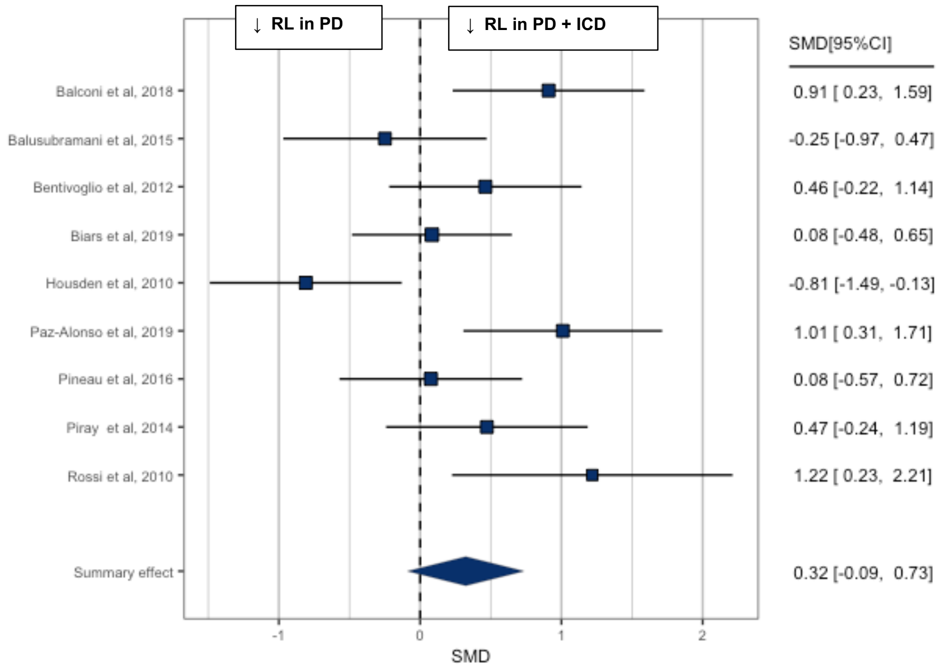

**Formulae used to convert study measures into Cohen's ds and associated variances.**

The following formulae were used to convert study measures into Cohen's ds and associated variances **between subjects:**

$$d = \frac{M_1 - M_2}{SD_{pooled}}$$

**Equation 1.** Cohen's d from Means and Standard Deviations of 2 samples. d is Cohen's d, M1 is the mean of one sample M2 is the mean of the other sample, SDpooled is the pooled standard deviation of the two samples (please see below.)

$$SD_{pooled} = \sqrt{(N_1 - 1)SD_1^2 + (N_2 - 1)SD_2^2}$$

**Equation 2.** Pooled standard deviation of 2 samples. SDpooled is the pooled standard deviation of the two samples, N1 is the size of one sample N2 is the size of the other sample, SD1 is the standard deviation of one sample, SD2 is the standard deviation of the other sample.

$$d = \frac{t}{\sqrt{\frac{1}{\frac{1}{N_1} + \frac{1}{N_2}}}}$$

**Equation 3.** Cohen's d from t-statistic. d is Cohen's d, N1 is the size of one sample N2 is the size of the other sample, t is the t-statistic

$$d = \frac{F}{\sqrt{\frac{1}{\frac{1}{N_1} + \frac{1}{N_2}}}}$$

**Equation 4.** Cohen's d from F-statistic. d is Cohen's d, N1 is the size of one sample N2 is the size of the other sample, F is the F-statistic.

$$Var_d = \frac{N_1 + N_2}{N_1 \times N_2} + \frac{d^2}{2(N_1 + N_2)}$$

**Equation 5.** Variance on Cohen's d for between subjects. Vard is the Variance on Cohen's d, N1 is the size of one sample N2 is the size of the other sample, d is Cohen's d.

The following formulae were used to convert study measures into Cohen's ds and associated variances **within subjects:**

$$d = \frac{M_d}{SD_d}$$

**Equation 6.** Cohen's d from Means and Standard Deviations from within subjects sample. Where $M_d$ is the mean change and $SD_d$ is the SD of the change scores
(equal to $SD_d = \sqrt{SD_1^2 + SD_2^2 - 2 \times r \times SD_1 SD_2}$ ).

$$d = \frac{t}{\sqrt{N}}$$

**Equation 7.** Cohen's d from t-statistic for within subjects results. d is Cohen's d, N is the sample size, t is the t-statistic

$$d = \sqrt{\frac{F}{N}}$$

**Equation 8.** Cohen's d from F-statistic. d is Cohen's d, N is the sample size, F is the F-statistic.

$$Var[d] = \frac{1}{n} + \frac{d^2}{2n}$$

**Equation 9.** Variance on Cohen's d for within subjects, n is the size of the sample.

### Supplement table 1. Option Valuation study characteristics and participant demographics.

| Option valuation in Parkinson's vs Healthy Controls | | | | Age (years) | | | Disease duration (years) | | UPDRS | | Cognition | | | Depression | | |
|---|---|---|---|---|---|---|---|---|---|---|---|---|---|---|---|---|
| Study | Task | Group | n | Mean | SD | % female | Mean | SD | Mean | SD | Cognitive measure | Mean | SD | Depression measure | Mean | SD |
| Bayard, 2016 | Game of dice task | PD | 78 | 67.49 | 8.16 | 35 | 7 | . | 24 | (5-80) | MMSE | 28.1 | 1.93 | BDI | 12.5 | 7.38 |
| | | HC | 96 | 67.95 | 6.75 | 32 | - | - | - | - | | 28.46 | 1.38 | | 10.04 | 11.76 |
| Brandt, 2015 | Game of dice task | PD | 15 | 64.78 | 8.09 | 46.66 | . | . | 14.43 | 10.14 | MoCA | 26.87 | 1.69 | GDS-15 | 3.33 | 3.54 |
| | | HC | 15 | 62.39 | 10.04 | 40 | - | - | - | - | | 27.6 | 1.43 | | 1.33 | 1.18 |
| Chong, 2015 | Effort based decision making task | PD | 26 | 66.6 | 6.8 | 34.6 | . | . | 21.6 | 11.7 | MoCA | 28.2 | 1.3 | DASS | 2 | 2.23 |
| | | HC | 26 | 66.2 | 6.4 | 42.3 | - | - | - | - | | 28.2 | 1.7 | | 1.5 | 1.84 |
| Cools, 2003 | Decision making gambling task | PD | 12 | 64.6 | 1.5 | 58.33 | 6.5 | 1.4 | 30.9 | 6.8 | MMSE | 29.5 | 0.15 | BDI | 7.7 | 1.1 |
| | | HC | 12 | . | . | . | - | - | - | - | | . | . | | . | . |
| Kobayashi, 2019 | Economic choice task | PD | 15 | 63 | 6.58 | 40 | 6.93 | 6.2 | 39 | 14.6 | MMSE | 28.3 | 1.76 | . | . | . |
| | | HC | 21 | 60 | 6.69 | 57.1 | - | - | - | - | | 28.7 | 1.99 | | . | . |
| Le Bouc, 2016 | Incentivised grip force choice task | PD | 24 | 60.2 | 1.6 | 29.17 | 11.4 | 1.3 | 11.7 | 1.7 | MMSE | 27.3 | 1.3 | MADRS | 5.9 | 1 |
| | | HC | 25 | 57 | 2.1 | 52 | - | - | - | - | | . | . | | 2.9 | 0.7 |
| Le Heron, 2018 | Effort based decision making task | PD | 39 | 67.8 | 7.6 | 19.04 | . | . | 29.5 | 9.9 | ACE | 91.1 | 7.9 | BDI | 14.3 | 7.7 |
| | | HC | 32 | 68.9 | 6.9 | 40.6 | - | - | - | - | | 95.6 | 3.8 | | 3.8 | 3.7 |
| Mcguigan, 2018 | Cognitive effort task | PD | 20 | 67.1 | 9.1 | 40 | 5.4 | 4.9 | 27.3 | 18.1 | MoCA | 27.7 | 1.92 | BDI | 9.5 | 4.98 |
| | | HC | 20 | 61.1 | 13.6 | 40 | - | - | - | - | | 28.1 | 1.37 | | 3.55 | 4.13 |
| Torta, 2009 | Cambridge gamble task | PD | 15 | 58.4 | 6.9 | 13.33 | 13.2 | 3.2 | 16.52 | 7.5 | MMSE | 28.2 | 1.5 | BDI | 10 | 6.6 |
| | | HC | 13 | 58.2 | 5.7 | 15.38 | - | - | - | - | | . | . | | . | . |
| Sharp, 2013 | Vancouver gambling task (modified) | PD | 18 | 65.47 | 9.17 | 27.78 | 5.59 | 4.04 | 20.65 | 6.91 | MOCA | 27.94 | 1.14 | BDI | 6.06 | 3.78 |
| | | HC | 18 | 66.76 | 5.83 | 50 | - | - | - | | | 28.85 | 1.32 | | 5.18 | 3.57 |
| **Option valuation in Parkinson's with psychiatric syndrome vs without** | | | | **Age (years)** | | | **Disease duration (years)** | | **UPDRS** | | **Cognition** | | | **Depression** | | |
| **Study** | **Task** | **Group** | **n** | **Mean** | **SD** | **% female** | **Mean** | **SD** | **Mean** | **SD** | **Cognitive measure** | **Mean** | **SD** | **Depression measure** | **Mean** | **SD** |
| Haagensen, 2020 | Game of dice task | PD | 13 | 61.4 | 9.7 | 46.15 | 4.5 | 2 | 22.8 | 6.9 | MoCA | 28.7 | 1.3 | BDI | 7 | 5 |
| | | ICD | 13 | 59.4 | 10.9 | 38.46 | 6.5 | 3.6 | 22.6 | 6 | | 28.7 | 0.9 | | 7.4 | 6.5 |
| Kobayashi, 2019 | Economic choice task | PD | 15 | 63 | 6.58 | 40 | 6.93 | 6.2 | 39 | 14.6 | MMSE | 28.3 | 1.76 | . | . | . |
| | | ICD | 10 | 63.1 | 9.68 | 40 | 8.4 | 2.99 | 49.9 | 26.3 | | 28.3 | 1.77 | . | . | . |
| Le Heron, 2018 | Effort based decision | PD | 18 | 68.2 | 6.5 | 44.44 | . | . | 27.1 | 13.2 | ACE | 93.5 | 5 | BDI | 11 | 7 |
| | | Apathy | 21 | 67.5 | 8.5 | 19.05 | . | . | 29.5 | 9.9 | | 89.4 | 9.4 | | 17.1 | 7.4 |

| | | | | | | | | | | | | | | | |
|---|---|---|---|---|---|---|---|---|---|---|---|---|---|---|---|
| Voon, 2011 | Gambling task | PD | 14 | 54.5 | 12.5 | 71.4 | . | . | . | . | . | . | . | . | . | . |
| | | ICD | 14 | 51.5 | 8.3 | 71.4 | . | . | . | . | . | . | . | . | . | . |

PD = Parkinson's disease, HC = healthy controls, ICD = Impulse control disorder, SD = standard deviation, IQR = interquartile range, '.' = not reported, '-' = not applicable, UPDRS = Unified Parkinson's Disease Rating Scale, MMSE = Mini-mental state examination, MoCA = Montreal cognitive assessment, ACE = Addenbrooke's cognitive examination, BDI = Beck depression inventory, DASS = Depression, Anxiety and Stress Scale, GDS = Geriatric depression scale, MADRS = Montgomery-Asberg Depression Rating Scale

### Supplement table 2. Reinforcement Learning study characteristics and participant demographics.

| Reinforcement learning in Parkinson's vs Healthy Controls | | | | Age (years) | | | Disease duration (years) | | UPDRS | | Cognition | | | Depression | | |
|---|---|---|---|---|---|---|---|---|---|---|---|---|---|---|---|---|
| Study | Task | Group | n | Mean | SD or (IQR) | % female | Mean | SD or (IQR) | Mean | SD or (IQR) | Measure | Mean | SD or (IQR) | Measure | Mean | SD or (IQR) |
| Castrioto, 2015 | IGT | PD | 20 | 53.2 | 6.6 | 45 | 10.3 | 3.8 | 12.3 | 6.1 | MDRS | 137.6 | 4.2 | . | . | . |
| | | HC | 24 | 54.9 | 7.6 | 62.5 | - | - | - | - | | 140.7 | 2.4 | | . | . |
| Balusubramani, 2015 | Probabilistic rewarded categorisation learning task | PD | 30 | . | . | 18.75 | 9.5 | . | . | . | MMSE | . | . | BDI | . | . |
| | | HC | 20 | . | . | 13.04 | - | - | - | - | | | | | . | . |
| Bodi, 2009 | Probabilistic classification task | PD | 22 | 44.8 | 5.2 | 30.77 | 0.266 | 0.166 | 27.5 | 6.1 | . | . | . | HAM-D | 4.2 | 1.4 |
| | | HC | 20 | 45.3 | 8.5 | 25 | - | - | - | - | | . | . | | . | . |
| Buelow, 2014 | IGT | PD | 24 | 68.04 | 7.86 | 45.8 | . | . | 29.89 | 13.14 | MMSE | 28.33 | 1.63 | GDS | 2.5 | 1.91 |
| | | HC | 14 | 69.62 | 6.36 | 53.85 | - | - | - | - | | 29.31 | 1.11 | | 1.15 | 1.57 |
| Cavanagh, 2017 | Cost of conflict task | PD | 28 | 69.75 | 8.59 | 39.28 | 5.54 | 4.18 | | | MMSE | 28.64 | 1.06 | BDI | 7.64 | 5.23 |
| | | HC | 28 | 69.21 | 9.23 | 39.28 | - | - | - | - | | 28.82 | 1.02 | | 4.93 | 4.69 |
| Czernecki, 2002 | IGT | PD | 23 | 57.6 | 2.1 | 60.87 | 14.9 | 1.2 | 12.4 | 2 | MDRS | 139.1 | 0.8 | MADRS | 8.3 | 1.4 |
| | | HC | 28 | 58.1 | 1.7 | 35.71 | - | - | - | - | | 141.1 | 0.3 | | 6.2 | 0.8 |
| Delazer, 2009 | IGT | PD | 20 | 68.5 | 5.9 | 25 | 5.25 | 6.38 | 17.6 | 8.7 | MMSE | 27.8 | 1.9 | HADS | 6.6 | 3.2 |
| | | HC | 20 | 71.3 | 3.5 | 85 | - | - | - | - | | 29.8 | 0.4 | | . | . |
| Euteneuer, 2009 | IGT | PD | 21 | 67.6 | 7.31 | 67 | 7.14 | 6.06 | 17.7 | 9.2 | MMSE | 29 | 1.1 | BDI | 3.9 | 2.12 |
| | | HC | 23 | 64.4 | 8.56 | 48 | | | | | | 29.65 | 0.65 | | 0.83 | 1.3 |
| Evens, 2015 | IGT | PD | 32 | 65.12 | 8.25 | 25 | 7.38 | 4.33 | 16.58 | 7.36 | MMSE | 29.25 | 0.98 | MADRS | 4.33 | 3.68 |
| | | HC | 32 | 65.53 | 5.94 | 31 | - | - | - | - | | 29.34 | 0.83 | | 1.59 | 2.19 |
| Garofalo, 2017 | Instrumental conditioning task | PD | 17 | 63.29 | 9.94 | 50 | 16.42 | 28.77 | . | . | MMSE | 28.41 | 1.37 | BDI | 12.66 | 7.83 |
| | | HC | 24 | 61.91 | 5.83 | 44 | - | - | - | - | | 28.94 | 1.54 | | 8.88 | 4.94 |
| Gescheidt, 2012 | IGT | PD | 19 (early onset) | 50.32 | 8.74 | 25 | 11.32 | 6.42 | 14.6 | 8.7 | MMSE | 29.37 | 0.96 | MADRS | . | . |
| | | HC | 20 | 49.95 | 9.03 | 26.3 | - | - | - | - | | 29.7 | 0.47 | | . | . |
| Graef, 2010 | | PD | 15 | 65.27 | 8.14 | 40 | 4.3 | 4 | 19.64 | 7.7 | MMSE | 29 | 1 | BDI | 6.93 | 5.55 |

| | | | | | | | | | | | | | | | | |
|---|---|---|---|---|---|---|---|---|---|---|---|---|---|---|---|---|
| | Probabilistic reversal learning task | HC | 16 | 67.75 | 4.55 | 37.5 | - | - | - | - | | 28.64 | 1.15 | | 5.31 | 2.98 |
| Herzallah, 2017 | Rewarded categorisation task | PD | 17 | 59.4 | 12.6 | 11.76 | 5.24 | 4 | 28.6 | 14.6 | MMSE | 28.5 | 1.1 | BDI | 8.3 | 5.2 |
| | | HC | 15 | 54.3 | 12.3 | 33.33 | - | - | - | - | | 29.3 | 0.7 | | 6.7 | 5.3 |
| Housden, 2010 | Rewarded salience attribution test | PD | 18 | 67.7 | 5.5 | 38.89 | 12.9 | 8.3 | 20 | 6.6 | MMSE | 28.6 | 2.1 | BDI | 12.9 | 9.9 |
| | | HC | 20 | 65.5 | 6 | 33.33 | - | - | - | - | | 29.4 | 0.8 | | 11.3 | 6.9 |
| Ibarretxe-Bilbao, 2009 | IGT | PD | 24 | 56.13 | 8.5 | 33 | 3.06 | 1.6 | 14.67 | 3.5 | MMSE | 29.63 | 0.5 | BDI | . | . |
| | | HC | 24 | 57.58 | 8.9 | 33 | - | - | - | - | | 29.83 | 0.4 | | | |
| Kobayakawa, 2008 | IGT | PD | 34 | 69.9 | 8.9 | 64.7 | 6.4 | 3.4 | . | . | MMSE | 28 | 2.2 | SDS | 36.1 | 8.9 |
| | | HC | 22 | 67.6 | 6.9 | 40.9 | - | - | - | - | | 28.8 | 1.6 | | 29.1 | 6 |
| Kobayakawa, 2010 | IGT | PD | 14 | 68.9 | 8 | 50 | 5.6 | 2.7 | . | . | MMSE | 28.2 | 1.9 | SDS | 32.9 | 7.8 |
| | | HC | 22 | 67.6 | 6.9 | 69.2 | - | - | - | - | | 28.8 | 1.6 | | 29.1 | 6 |
| Mapelli, 2014 | IGT | PD | 15 | 61.4 | 9.6 | 25 | 4.8 | 3.4 | 8.9 | 4 | MMSE | 28.3 | 1.2 | BDI | Excluded if >14 | |
| | | HC | 15 | 60.7 | 9.8 | 26.7 | - | - | - | - | | 27.86 | 1.5 | | . | . |
| Mimura, 2006 | IGT | PD | 18 | 68.9 | 7 | 72.2 | . | . | . | . | MMSE | 27.8 | 1.9 | ZSRD | 39.4 | 6.9 |
| | | HC | 20 | . | . | 70 | - | - | - | - | | 29.1 | 1.5 | | 30.5 | 6 |
| Pagonabarraga, 2007 | IGT | PD | 35 | 67.2 | 8 | 37.2 | 8.4 | 5 | 21.2 | 8 | MDRS | 133 | 6 | . | . | . |
| | | HC | 31 | 70.2 | 10 | 47.7 | - | - | - | - | | 136 | 5 | | . | . |
| Paz-Alonso, 2019 | IGT | PD | 17 | 61 | 8.7 | 11.8 | 7 | (4-10) | 25.9 | 8.2 | . | . | . | HADS-D | 2.2 | 2.6 |
| | | HC | 18 | 63 | 9.7 | 16.7 | - | - | - | - | | . | . | | 1.9 | 1.3 |
| Perretta, 2005 | IGT | PD (late stage) | 16 | 77.7 | 6 | 50 | . | . | 27.2 | 1.3 | MMSE | Excluded if <27 | . | BDI | 12.7 | 1.5 |
| | | PD (early stage) | 16 | 72.4 | 2.3 | 43.75 | . | . | 11.3 | 1.1 | | | | | 6.9 | 0.7 |
| | | HC | 19 | 72.6 | 8.28 | 42.1 | - | - | - | - | | | | | 5.2 | 0.8 |
| Pignatti, 2012 | IGT | PD | 15 | 64.27 | 10.5 | . | . | . | . | . | MMSE | 28.31 | 1.11 | . | . | . |
| | | HC | 16 | 41.56 | 13.9 | . | - | - | - | - | | | | | . | . |
| Piray, 2014 | Rewarded categorisation task | PD | 40 | 63.33 | 3.98 | 23.07 | 9.72 | 2.64 | 19.6 | 6.42 | MMSE | 27 | 0.93 | BDI | 8 | 1.69 |
| | | HC | 20 | 66.45 | 4.7 | 35 | - | - | - | - | | 27.65 | 1.18 | | 7.75 | 1.97 |
| Poletti, 2010 | IGT | PD (off) | 24 | 64.9 | 5.8 | 27 | 0 (de novo) | | 14.3 | 8.1 | MMSE | 28.8 | 2 | GDS | 4.7 | 3.3 |
| | | HC | 25 | 65.4 | 2.2 | 44 | | | | | | 28.5 | 1.8 | | 2.8 | 1.4 |
| Thiel, 2003 | IGT | PD | 5 | 62.6 | 12.5 | 40 | 8 | 2.55 | 20.6 | 6.47 | MMSE | .28.4 | 2.07 | . | . | . |
| | | HC | 5 | 44.2 | 21.3 | 20 | - | - | - | - | | | | | | |
| Xi, 2015 | IGT | PD | 15 | 60.73 | 11.7 | 53 | 4.33 | 5.05 | 15.87 | 8.96 | MMSE | 27.6 | 4.63 | HAMD | 4.67 | 1.29 |

| | | | | | | | | | | | | | | | | |
|---|---|---|---|---|---|---|---|---|---|---|---|---|---|---|---|---|
| | | HC | 15 | 56.33 | 14.5 | 60 | - | - | - | - | | 29.4 | 1.12 | | 3.8 | 1.37 |
| Yildirim, 2020 | IGT | PD | 39 | 65.9 | 7.3 | 21 | 7.8 | 4.3 | 11.6 | 6.1 | MMSE | . | . | . | . | . |
| | | HC | 37 | 64.4 | 8 | 24 | - | - | - | - | | . | . | | . | . |
| **Reinforcement learning in Parkinson's with psychiatric syndrome vs without** | | | | **Age** | | | **Disease duration (years)** | | **UPDRS** | | **Cognition** | | | | **Depression** | |
| **Study** | **Task** | **Group** | **n** | **Mean** | **SD or (IQR)** | **% female** | **Mean** | **SD or (IQR)** | **Mean** | **SD or (IQR)** | **Measure** | **Mean** | **SD or (IQR)** | **Depression measure** | **Mean** | **SD or (IQR)** |
| Balconi, 2018 | IGT | PD | 20 | 63.9 | 7.1 | 15 | . | . | 13 | 9.3 | . | . | . | BDI | 11.3 | 6 |
| | | PG | 17 | 60.7 | 6.1 | 17.65 | . | . | 17 | 7.8 | | . | . | | 15.9 | 9.1 |
| Balusubramani, 2015 | Rewarded categorisation task | PD | 14 | . | . | 21.4 | 8.35 | . | . | . | MMSE | . | . | BDI | . | . |
| | | ICD | 16 | . | . | 12.5 | 9.56 | . | | | | | | | | |
| Bentivoglio, 2012 | IGT | PD | 17 | 63.94 | 9.2 | 35.29 | 7.3 | 4.4 | 22.5 | 6.9 | MMSE | 28.6 | 1.4 | HAM-D | 5.6 | 4.1 |
| | | ICD | 17 | 62 | 10.1 | 17.65 | 6.9 | 3.8 | 23.8 | 11 | | 28.4 | 1.6 | | 7.5 | 6.3 |
| Biars, 2019 | IGT | PD | 24 | 60.5 | 7.8 | 12.5 | 11.9 | 7.1 | 37.2 | 14.8 | DRS | 136.5 | 5.9 | BDI | 8.5 | 5.3 |
| | | ICD | 24 | 61.2 | 8.3 | 12.5 | 13.2 | 7.1 | 21.6 | 9.9 | | 138 | 3.5 | | 11.2 | 6.8 |
| Buelow, 2014 | IGT | PD | 14 | 69 | 6.19 | 57.14 | . | . | 29.62 | 6.85 | MMSE | 28.71 | 1.38 | GDS | 2.14 | 1.41 |
| | | Apathy | 10 | 66.7 | 9.96 | 30 | . | . | 29.89 | 13.14 | | 27.8 | 1.87 | | 3 | 2.45 |
| Garofalo, 2017 | Instrumental conditioning task | PD | 17 | 63.29 | 9.94 | 44 | 10.94 | 10.38 | . | . | MMSE | 28.94 | 1.54 | BDI | 8.88 | 4.94 |
| | | Psychosis | 12 | 60.83 | 6.6 | 50 | 16.42 | 28.77 | . | . | | 28.41 | 1.37 | | 12.66 | 7.83 |
| Herzallah, 2017 | Rewarded categorisation task | PD | 17 | 59.4 | 12.6 | 11.76 | 5.24 | 4 | . | . | MMSE | 28.5 | 1.1 | BDI | 8.3 | 5.2 |
| | | MDD | 13 | 55.2 | 11.9 | 69.2 | 4.84 | 3.7 | 28.6 | 14.6 | | 27.5 | 1.8 | | 26.9 | 7.7 |
| Housden, 2010 | Rewarded salience attribution test | PD | 18 | 67.7 | 5.5 | 33.33 | 12.9 | 8.3 | 21.3 | 0.4 | MMSE | 29.4 | 0.8 | BDI | 11.3 | 6.9 |
| | | ICD | 18 | 62.3 | 7.6 | 38.89 | 13.9 | 9 | 20 | 6.6 | | 28.6 | 2.1 | | 12.9 | 9.9 |
| Martinez-Horta, 2013 | IGT | PD | 17 | 65.06 | 4.85 | . | 5.38 | 4.25 | 18 | 4.61 | MMSE | 28.94 | 1.24 | HADS | 3.88 | 3.6 |
| | | Apathy | 17 | 68.25 | 6.06 | . | 6.75 | 4.93 | 18.65 | 5.8 | | 28.55 | 1.23 | | 5.65 | 2.92 |
| Paz-Alonso, 2019 | IGT | PD | 17 | 61 | 8.7 | 11.8 | 7 | (4-10) | 25.9 | 8.2 | . | . | . | HADS | 2.2 | 2.6 |
| | | ICD | 18 | 62.3 | 7.6 | 11.1 | 8 | (5.1-10) | 22.31 | 6.6 | | . | . | | 3.1 | 2.4 |
| Pineau, 2016 | IGT (adapted) | PD | 20 | 55 | (40-62) | 35 | 5.5 | (4-12) | 8.5 | (0-34) | MDRS | 139 | (131-143) | . | . | |
| | | ICD | 17 | 55 | (37-65) | 17.64 | 7 | (2-10) | 7 | (0-23) | | 140 | (133-144) | | . | . |
| Piray, 2014 | Rewarded categorisation task | PD | 40 | 63.33 | 3.98 | 25.9 | 8.87 | 3.14 | 19.6 | 6.42 | MMSE | 27 | 0.93 | BDI | 8 | 1.69 |
| | | ICD | 16 | 64.38 | 3.32 | 11.1 | 9.63 | 2.45 | 19 | 5.32 | | 27.19 | 1.11 | | 6.75 | 1.69 |
| Poletti, 2011 | IGT | PD | 12 | 63.92 | 7.17 | 16.66 | . | . | . | . | MMSE | 28.72 | 2.41 | GDS-15 | 3.33 | 2.34 |
| | | Alexithymia | 12 | 66.17 | 5.2 | 41.66 | . | . | . | . | | 28.49 | 1.94 | | 6.33 | 3.89 |
| Rossi, 2010 | IGT | PD | 13 | 65.1 | 3.8 | 23 | . | . | 14.7 | 6.7 | MMSE | . | . | MADRS | 14.1 | 7.9 |

| Study | Task | Group | n | Age Mean | Age SD | % female | Disease duration Mean | Disease duration SD | UPDRS Mean | UPDRS SD | Cognitive measure | Cognition Mean | Cognition SD | Depression measure | Depression Mean | Depression SD |
|---|---|---|---|---|---|---|---|---|---|---|---|---|---|---|---|---|
| | | PG | 7 | 61.4 | 6.9 | 14 | . | . | 17 | 9.1 | | Excluded if <24 | . | | 17.1 | 6.5 |
| Sáez-Francàs, 2014 | IGT | PD | 56 | 62.64 | 9.06 | 32.14 | 4.31 | 3.65 | 18.41 | 5.82 | MMSE | . | . | HAM-D | 3.64 | 3.37 |
| | | Fatigue | 33 | 61.73 | 9.85 | 39.4 | 4.94 | 3.44 | 20.85 | 6.31 | | Excluded if <26 | . | | 8.64 | 7.06 |
| Timmer, 2017 | Rewarded task-switching paradigm | PD | 22 | 61.1 | 7.6 | 36.4 | . | . | 21.9 | 6.8 | MMSE | 28.6 | 1.2 | BDI | 4.3 | 2.3 |
| | | MDD | 19 | 58.4 | 5.3 | 36.8 | . | . | . | . | | 28.5 | 1.3 | | 8.7 | 5 |

PD = Parkinson's disease, HC = healthy controls, IGT = Iowa Gambling Task, ICD = Impulse control disorder, MDD = Major depressive disorder, SD = standard deviation, IQR = interquartile range, '.' = not reported, '-' = not applicable, UPDRS = Unified Parkinson's Disease Rating Scale, MMSE = Mini-mental state examination, MoCA = Montreal cognitive assessment, MDRS = Mattis dementia rating scale, BDI = Beck depression inventory, HAM-D = Hamilton depression rating scale, HADS = Hospital anxiety and depression scale ZSRD = Zung Self-Rating Depression scale, DASS = Depression, Anxiety and Stress Scale, GDS = Geriatric depression scale, MADRS = Montgomery-Asberg Depression Rating Scale, SDS = Self-rated depression scale

### Supplement table 3. Response Vigor and Reward Bias study characteristics and participant demographics.

| Response vigor in Parkinson's vs Healthy Controls | | | | Age | | | Disease duration | | UPDRS | | Cognition | | | Depression | | |
|---|---|---|---|---|---|---|---|---|---|---|---|---|---|---|---|---|
| **Study** | **Task** | **Group** | **n** | **Mean** | **SD** | **% female** | **Mean** | **SD** | **Mean** | **SD** | **Cognitive measure** | **Mean** | **SD** | **Depression measure** | **Mean** | **SD** |
| Muhammed, 2016 | Speed of gaze shifting task | PD | 16 | 67.3 | 6.4 | . | 3.2 | 2.2 | 19.4 | 9.8 | MoCA | 27.8 | 2.3 | BDI | 11.7 | 5.5 |
| | | HC | 31 | 65.9 | 5.6 | . | - | - | - | - | | 28.2 | 1.6 | | 15.2 | 7.8 |
| Renfroe, 2016 | Rewarded speed of response to specific visual stimuli | PD | 18 | 66 | 7.99 | 22.2 | 8.39 | 4.57 | 26 | 9.55 | . | . | . | BDI | 7.82 | 4.85 |
| | | HC | 15 | 70 | 6.94 | 40 | - | - | - | - | | . | . | | 1.8 | 2.44 |
| Timmer, 2018 | Stroop-like incentive task switching | PD | 23 | 61 | 7.4 | 39.13 | 4.5 | 2.2 | 21.8 | 6.7 | MMSE | 28.5 | 1.3 | BDI | 4.1 | 2.3 |
| | | HC | 23 | 60.9 | 5.9 | 39.13 | - | - | - | - | | 28.8 | 1.2 | | 3.1 | 2.1 |
| **Response vigor in Parkinson's with psychiatric syndrome vs without** | | | | **Age** | | | **Disease duration** | | **UPDRS** | | **Cognition** | | | **Depression** | | |
| **Study** | **Task** | **Group** | **n** | **Mean** | **SD** | **% female** | **Mean** | **SD** | **Mean** | **SD** | **Cognitive measure** | **Mean** | **SD** | **Depression measure** | **Mean** | **SD** |
| Drew, 2020 | Speed of gaze shifting task | PD | 26 | 67.19 | 5.92 | 26.9 | 4.87 | 4.09 | 18.62 | 9.38 | MoCA | 27.77 | 1.95 | BDI | 13 | 7.1 |
| | | ICD | 23 | 63.7 | 7.56 | 47.82 | 8.71 | 4.25 | 24.22 | 16.94 | | 27.39 | 2.55 | | 12.26 | 5.84 |
| Evans, 2010 | Card arranging reward responsivity test | PD | 20 | 59.5 | 7.9 | . | 13.6 | 8 | 19.2 | 2.5 | MMSE | 29.1 | (27-30) | GDS | 9 | 4.9 |
| | | DDS | 20 | 55.4 | 7.6 | . | 14 | 5.7 | 22.7 | 2.5 | | 29 | (24-30) | | 17.6 | 6.4 |
| Lawrence, 2011 | Spatial search task | PD | 10 | 59.6 | 6.4 | . | . | . | 25.6 | 11 | MMPD | 25.6 | 11 | GDS | 4.7 | 2 |
| | | Apathy | 10 | 61.7 | 6.1 | . | . | . | 28.2 | 11.4 | | 29.3 | 2.8 | | 8.2 | 3.5 |
| Muhammed, 2016 | Speed of gaze shifting task | PD | 16 | 65.9 | 5.6 | . | 6.8 | 4.6 | 18.4 | 2.6 | MoCA | 28.2 | 1.6 | BDI | 11.7 | 5.5 |
| | | Apathy | 14 | 67.3 | 6.4 | . | 3.2 | 2.2 | 20.4 | 2.4 | | 27.8 | 2.3 | | 15.2 | 7.8 |
| Timmer, 2018 | Stroop-like incentive task switching | PD | 23 | 61 | 7.4 | 39.13 | 4.5 | 2.2 | 21.8 | 6.7 | MMSE | 28.5 | 1.3 | BDI | 4.1 | 2.3 |
| | | Depression | 22 | 58.4 | 5.7 | 36.36 | 5 | 3.5 | 23.1 | 9.6 | | 28.4 | 1.4 | | 9.6 | 6.1 |

PD = Parkinson's disease, HC = healthy controls, ICD = Impulse control disorder, SD = standard deviation, IQR = interquartile range, '.' = not reported, '-' = not applicable, UPDRS = Unified Parkinson's Disease Rating Scale, MMSE = Mini-mental state examination, MoCA = Montreal cognitive assessment, MMPD = Mini-mental Parkinson's examination BDI = Beck depression inventory, GDS = Geriatric depression scale

### Supplement table 4. Study summary statistics, tasks and measures included in the Meta-Analysis of Parkinson's versus healthy controls.

| Reward processing summary statistics for Parkinson's disease patients versus healthy controls | | | | | | | | | | | | | Tasks and Measures Included in the Meta-Analysis. | |
|---|---|---|---|---|---|---|---|---|---|---|---|---|---|---|
| Author, Year | Category | OFF/ON | N (HC) | N (PD) | t | F | M (HC) | SD (HC) | M (PD) | SD (PD) | d | Vard | Task | Measure |
| Bayard et al, 2016 (ON) | OV | ON | 96 | 78 | | | 6.90 | 7.65 | 2.77 | 8.41 | 0.52 | 0.02 | Game of dice task | Net score |
| Chong et al, 2015 (ON) | OV | ON | 26 | 26 | | | | | | | -0.35 | 0.08 | Effort based decision making task (apple gathering) | Effort indifference point (mean effect size across stake levels, fig 4) |
| Chong et al, 2015 (OFF) | OV | OFF | 26 | 26 | | 2.70 | | | | | 0.46 | 0.08 | Effort based decision making task (apple gathering) | Effort indifference point |
| Brandt et al, 2015 (ON) | OV | ON | 15 | 15 | | | 14.93 | 15.72 | -8.67 | 17.67 | 1.41 | 0.17 | Game of dice task | Net score (fig 1) |
| Kobayashi et al, 2019 (ON) | OV | ON | 21 | 15 | | | 0.21 | 0.78 | 0.30 | 0.66 | 0.12 | 0.11 | Economic choice task | Relative risk aversion coefficient (fig 2C, first session) |
| Kobayashi et al, 2019 (OFF) | OV | OFF | 21 | 15 | | | 0.21 | 0.78 | 0.97 | 0.70 | 1.02 | 0.13 | Economic choice task | Relative risk aversion coefficient (fig 2C, first session) |
| Le Heron et al, 2018 (ON) | OV | ON | 32 | 18 | 1.30 | | | | | | 0.38 | 0.09 | Effort based decision making task (apple gathering) | Mean difference in proportion of offers accepted |
| Mcguigan et al, 2018 (ON) | OV | ON | 20 | 20 | -0.32 | | | | | | -0.10 | 0.10 | Cognitive effort task | Mean difference in k-value |
| Mcguigan et al, 2018 (OFF) | OV | OFF | 20 | 20 | 2.79 | | | | | | 0.88 | 0.11 | Cognitive effort task | Mean difference in k-value |
| Sharp et al, 2013 (ON) | OV | ON | 18 | 18 | 0.43 | | | | | | 0.14 | 0.07 | Vancouver gambling task | Gain phase adjust y intercept |
| Sharp et al, 2013 (OFF) | OV | OFF | 18 | 18 | 1.79 | | | | | | 0.60 | 0.07 | Vancouver gambling task | Gain phase adjust y intercept |
| Torta et al, 2009 (ON) | OV | ON | 13 | 15 | | | | | | | 0.24 | 0.14 | Cambridge gamble task | Average bet (across risk levels) |
| Torta et al, 2009 (OFF) | OV | OFF | 13 | 15 | | | | | | | 0.23 | 0.14 | Cambridge gamble task | Average bet (across risk levels) |
| Cools et al, 2003 (ON) | OV | ON | 12 | 12 | | | 54.91 | 12.09 | 59.65 | 16.11 | -0.33 | 0.17 | Incentivised decision making task | Mean % bets across ascending & descending conditions |
| Cools et al, 2003 (OFF) | OV | OFF | 12 | 12 | | | 54.91 | 12.09 | 58.95 | 15.73 | -0.29 | 0.17 | Incentivised decision making task | Mean % bets across ascending & descending conditions |
| Le Bouc et al, 2016 (ON) | OV | ON | 25 | 24 | | | 80.89 | 7.75 | 78.56 | 12.49 | 0.23 | 0.08 | Effort based decision making task: Binary choice task | Choice of effort level at the highest stake level. (fig 4D) |
| Le Bouc et al, 2016 (OFF) | OV | OFF | 25 | 24 | 3.51 | | | | | | 1.00 | 0.09 | Effort based decision making task: Binary choice task | Choice of effort level at the highest stake level. (fig 4D) |
| Castrioto et al, 2015 (ON) | RL | ON | 24 | 20 | | | 4.38 | 40.22 | -1.79 | 44.72 | 0.15 | 0.09 | Iowa Gambling Task | Baseline final round mean score. (fig 1A) |
| Castrioto et al, 2015 (OFF) | RL | OFF | 24 | 20 | | | 4.38 | 40.22 | 0.98 | 40.34 | 0.08 | 0.09 | Iowa Gambling Task | Baseline final round mean score. (fig 1A) |
| Balusubramani et al, 2015 (ON) | RL | ON | 20 | 14 | | | 64.10 | 84.84 | 61.54 | 78.69 | 0.03 | 0.12 | Probabilistic rewarded categorisation learning task | % Optimality during expected reward (fig 3A) |
| Balusubramani et al, 2015 (OFF) | RL | OFF | 20 | 26 | | | 64.10 | 84.84 | 43.07 | 99.38 | 0.23 | 0.09 | Probabilistic rewarded categorisation learning task | % Optimality during expected reward (fig 3A) |
| Bodi et al, 2009 (ON) | RL | Medicated | 20 | 22 | | | 81.18 | 18.43 | 89.18 | 17.12 | -0.45 | 0.10 | Feedback-based probabilistic classification task | Final block % optimal decisions (fig 2) |
| Bodi et al, 2009 (Never medicated) | RL | Never medicated | 20 | 26 | | | 81.18 | 18.43 | 59.53 | 18.61 | 1.17 | 0.10 | Feedback-based probabilistic classification task | Final block % optimal decisions (fig 2) |
| Buelow et al, 2014 (ON) | RL | ON | 14 | 24 | | | | | | | 1.38 | 0.14 | Iowa Gambling Task | Final block score (authors provided) |

| | | | | | | | | | | | | | | |
|---|---|---|---|---|---|---|---|---|---|---|---|---|---|---|
| Piray et al, 2014 (ON) | RL | ON | 20 | 15 | | | 0.12 | 0.16 | 0.14 | 0.20 | -0.10 | 0.12 | Probabilistic learning task | Actor's learning rate (fig 6C) |
| Piray et al, 2014 (OFF) | RL | OFF | 20 | 25 | | | 0.12 | 0.16 | 0.08 | 0.13 | 0.29 | 0.09 | Probabilistic learning task | Actor's learning rate (fig 6C) |
| Yildirim et al, 2020 (ON) | RL | ON | 37 | 39 | | | 0.65 | 5.72 | -0.11 | 5.77 | 0.13 | 0.05 | Iowa Gambling Task | Final block mean score (author provided) |
| Gescheidt et al, 2012 (ON) | RL | ON | 20 | 19 | | | 10.30 | 29.42 | -6.00 | 25.26 | 0.59 | 0.11 | Iowa Gambling Task | Mean total score |
| Herzallah et al, 2017 (ON) | RL | ON | 15 | 17 | | | 76.49 | 17.36 | 72.10 | 16.62 | 0.26 | 0.13 | Feedback-based probabilistic classification task | Mean % optimal responses for positive feedback (fig 5A) |
| Housden et al, 2010 (ON) | RL | ON | 20 | 18 | | | 56.50 | 22.40 | 27.00 | 19.10 | 1.41 | 0.13 | Salience Attribution Test | Visual analogue scale rating for high probability stimuli |
| Mapelli et al, 2014 (ON) | RL | ON | 15 | 15 | | | 9.81 | 10.19 | 4.74 | 5.11 | 0.63 | 0.14 | Iowa Gambling Task | Final block mean score (fig 1) |
| Mimura et al, 2006 (ON) | RL | ON | 20 | 18 | | | 3.70 | 7.35 | -0.33 | 6.59 | 0.58 | 0.11 | Iowa Gambling Task | Final 50 cards mean score |
| Pagonabarraga et al, 2007 (ON) | RL | ON | 31 | 35 | | | 6.10 | 17.00 | -10.80 | 19.00 | 0.93 | 0.07 | Iowa Gambling Task | Mean total score |
| Paz-Alonso et al, 2019 (ON) | RL | ON | 18 | 17 | | | 40.60 | 18.82 | 41.80 | 15.38 | -0.07 | 0.11 | Iowa Gambling Task | Mean difference % optimal choice, final block. (supplement fig 1) |
| Garofalo et al, 2017 (ON) | RL | ON | 24 | 17 | | | 21.50 | 5.26 | 15.15 | 6.68 | 1.08 | 0.11 | Instrumental conditioning task | Reward learning index (fig 2B) |
| Czernecki et al, 2002 (ON) | RL | ON | 28 | 23 | | | 8.32 | 11.06 | 2.86 | 16.59 | 0.40 | 0.08 | Iowa Gambling Task | Final block mean score (fig 1, second session) |
| Czernecki et al, 2002 (OFF) | RL | OFF | 28 | 23 | | | 8.32 | 11.06 | 5.36 | 10.89 | 0.27 | 0.08 | Iowa Gambling Task | Final block mean score (fig 1, second session) |
| Graef et al, 2010 (ON) | RL | ON | 15 | 14 | 1.80 | | 68.49 | 14.07 | 60.07 | 11.08 | 0.67 | 0.15 | Instrumental learning task | % correct choices with constant reward contingencies |
| Graef et al, 2010 (OFF) | RL | OFF | 15 | 14 | 2.19 | | 68.49 | 14.07 | 59.05 | 8.86 | 0.81 | 0.15 | Instrumental learning task | % correct choices with constant reward contingencies |
| Thiel et al, 2003 (OFF) | RL | OFF | 5 | 5 | | | 53.6 | 15.8 | 61.2 | 17.1 | -0.46 | 0.41 | Iowa Gambling Task | Mean number of advantageous cards selected. |
| Perretta et al, 2005 (late PD) | RL | ON late PD | 19 | 16 | | | 7.80 | 1.31 | 6.80 | 1.60 | 0.69 | 0.12 | Iowa Gambling Task | Mean total score |
| Perretta et al, 2005 (early PD) | RL | ON early PD | 19 | 16 | | | 7.80 | 1.31 | 7.60 | 1.60 | 0.14 | 0.12 | Iowa Gambling Task | Mean total score |
| Kobayakawa et al, 2008 (ON) | RL | ON | 22 | 34 | | 4.80 | 4.90 | 12.20 | -16.00 | 21.57 | 0.60 | 0.08 | Iowa Gambling Task | Group difference in choice patterns (advantageous - disadvantageous) |
| Kobayakawa et al, 2010 (ON) | RL | ON | 22 | 14 | | 2.96 | | | | | 0.59 | 0.12 | Iowa Gambling Task | Group difference in choice patterns (advantageous - disadvantageous) |
| Poletti et al, 2010 (OFF) | RL | OFF (de novo) | 25 | 30 | | | 6.00 | 6.82 | 3.07 | 12.07 | 0.29 | 0.07 | Iowa Gambling Task | Mean total score |
| Delazer et al, 2009 (ON) | RL | ON | 20 | 20 | | 8.81 | | | | | 0.94 | 0.11 | Iowa Gambling Task | Group difference in choice patterns (advantageous - disadvantageous) |
| Euteneuer et al, 2009 (ON) | RL | ON | 23 | 21 | | 0.65 | | | | | 0.24 | 0.09 | Iowa Gambling Task | Group difference in choice patterns (advantageous - disadvantageous) |
| Ibarretxe-Bilbao et al, 2009 (ON) | RL | ON | 24 | 24 | | 14.56 | | | | | 1.10 | 0.10 | Iowa Gambling Task | Group difference in choice patterns (advantageous - disadvantageous) |
| Pignatti et al, 2012 (ON) | RL | ON | 16 | 15 | | | 12.50 | 14.87 | 7.47 | 16.06 | 0.33 | 0.13 | Iowa Gambling Task | Total score over the last 50 choices |
| Evens et al, 2015 (ON) | RL | ON | 32 | 32 | | | -7.31 | 20.78 | -2.09 | 16.27 | -0.28 | 0.06 | Iowa Gambling Task | Mean total score |

| | | | | | | | | | | | | | | |
|---|---|---|---|---|---|---|---|---|---|---|---|---|---|---|
| Xi et al, 2015 (ON) | RL | ON | 15 | 15 | | 10.67 | | | | | 1.19 | 0.16 | Iowa Gambling Task | Group difference in choice patterns (advantageous - disadvantageous) |
| Cavanagh et al, 2017 (ON) | RL | ON | 28 | 28 | -1.64 | | | | | | -0.44 | 0.07 | Cost of conflict task | % selection of most (A) vs least (D) rewarding stimuli |
| Cavanagh et al, 2017 (OFF) | RL | OFF | 28 | 28 | -1.44 | | | | | | -0.38 | 0.07 | Cost of conflict task | % selection of most (A) vs least (D) rewarding stimuli |
| Muhammed et al, 2016 (ON) | RV | ON | 31 | 30 | 5.50 | | | | | | 0.60 | 0.07 | Rewarded speed of gaze shifting task | Reward sensitivity in peak velocity |
| Timmer et al, 2018 (ON) | RV | ON | 23 | 23 | | | 5.00 | 4.50 | 14.40 | 4.40 | -2.11 | 0.14 | Rewarded task switching paradigm | Reward related speeding 'repeat' condition |
| Timmer et al, 2018 (OFF) | RV | OFF | 23 | 23 | | | 5.00 | 4.50 | 3.80 | 4.70 | 0.26 | 0.09 | Rewarded task switching paradigm | Reward related speeding 'repeat' condition |

### Supplement table 5. Study summary statistics, tasks and measures included in the Meta-Analysis of Parkinson's with & without psychiatric syndrome

| **Reward processing summary statistics for Parkinson's disease versus Parkinson's disease plus psychiatric syndrome** | | | | | | | | | | | | | **Tasks and Measures Included in the Meta-Analysis.** | |
|---|---|---|---|---|---|---|---|---|---|---|---|---|---|---|
| Author, Year | Category | Psychiatric syndrome | N (PD) | N (PD+PSYCH) | t | F | M (PD) | SD (PD) | M (PD +Psych) | SD (PD+Psych) | d | Vard | Task | Measure |
| Balusubramani et al, 2015 (ON) | RL | ICD | 14 | 16 | | | 61.54 | 78.69 | 78.97 | 61.56 | -0.25 | 0.13 | Probabilistic reward and punishment learning task | % Optimality during expected reward (fig 3A) |
| Piray et al, 2014 (ON) | RL | ICD | 15 | 16 | | | 0.135 | 0.198 | 0.067 | 0.06 | 0.47 | 0.13 | Probabilistic learning task | Actor's learning rate (fig 6C) |
| Housden et al, 2010 (ON) | RL | ICD | 18 | 18 | | | 27 | 19.1 | 42.7 | 19.7 | -0.81 | 0.12 | Salience Attribution Test | Visual analogue scale rating for high probability stimuli |
| Biars et al, 2019 (ON) | RL | ICD | 24 | 24 | | | 3.5 | 9.7 | 2.75 | 8.2 | 0.08 | 0.08 | Iowa Gambling Task | Final block score (authors provided) |
| Balconi et al, 2018 (ON) | RL | PG | 20 | 17 | | 7.60 | | | | | 0.91 | 0.12 | Iowa Gambling Task | Group difference in choice patterns (advantageous - disadvantageous) |
| Paz-Alonso et al, 2019 (ON) | RL | ICD | 17 | 18 | | | 41.80 | 15.38 | 27.00 | 13.94 | 1.01 | 0.13 | Iowa Gambling Task | Mean difference % optimal choice, final block. (supplement fig 1) |
| Pineau et al, 2016 (ON) | RL | ICD | 20 | 17 | | | 15.00 | 15.56 | 14.00 | 9.63 | 0.08 | 0.11 | Iowa Gambling Task | Mean number of cards selected from winning deck |
| Bentivoglio et al, 2012 (ON) | RL | ICD | 17 | 17 | | | 8.40 | 22.10 | -4.60 | 33.10 | 0.46 | 0.12 | Iowa Gambling Task | Mean total score |
| Rossi et al, 2010 (ON) | RL | PG | 13 | 7 | 2.60 | | | | | | 1.22 | 0.26 | Iowa Gambling Task | Final block mean net score |
| Garofalo et al, 2017 (ON) | RL | Psychosis | 17 | 12 | | | 15.15 | 6.68 | 8.67 | 8.34 | 0.88 | 0.16 | Instrumental conditioning task | Reward learning index (fig 2B) |
| Sáez-Francàs et al, 2014 (ON) | RL | Fatigue | 56 | 33 | | | 1.79 | 14.80 | -4.18 | 11.13 | 0.44 | 0.05 | Iowa Gambling Task | Final three blocks mean net score |
| Martinez-Horta et al, 2013 (ON) | RL | Apathy | 17 | 20 | | | -1.82 | 5.80 | 4.10 | 4.80 | -1.12 | 0.13 | Iowa Gambling Task | Final block score |
| Buelow et al, 2014 (ON) | RL | Apathy | 14 | 10 | | | 22.12 | 16.80 | -24.00 | 14.87 | 2.88 | 0.34 | Iowa Gambling Task | Final block score (authors provided) |
| Herzallah et al, 2017 (ON) | RL | MDD | 17 | 13 | | | 72.10 | 16.62 | 49.30 | 22.14 | 1.19 | 0.16 | Feedback-based probabilistic classification task | Mean % optimal responses for positive feedback (fig 5A) |
| Timmer et al, 2017 (ON) | RL | MDD | 22 | 19 | | | 0.10 | 0.06 | 0.07 | 0.06 | -0.50 | 0.10 | Deterministic reversal learning paradigm | Error rate for expected reward |

| | | | | | | | | | | | | | | |
|---|---|---|---|---|---|---|---|---|---|---|---|---|---|---|
| Poletti et al, 2011 (OFF) | RL | Alexithymia | 12 | 12 | | | 4.17 | 8.37 | 3.00 | 6.35 | 0.16 | 0.17 | Iowa Gambling Task | Final block score |
| Kobayashi et al, 2019 (ON) | OV | ICD | 15 | 10 | | | 0.30 | 0.66 | -0.19 | 0.17 | -0.93 | 0.18 | Economic choice task | Relative risk aversion coefficient (fig 2C, first session) |
| Le Heron et al, 2018 (ON) | OV | Apathy | 18 | 21 | 2.33 | | | | | | 0.75 | 0.11 | Effort based decision making task (apple gathering) | Mean difference in proportion of offers accepted |
| Voon et al, 2011 (ON) | OV | ICD | 14 | 14 | | | | | | | -0.68 | 0.14 | Gambling task | Proportion of risky choices in gain phase (across risk levels, fig 2A) |
| Timmer et al, 2018 (ON) | RV | MDD | 23 | 22 | | | 14.40 | 4.40 | 17.70 | 3.90 | -0.79 | 0.10 | Rewarded task-switching paradigm | Reaction time reward benefit (repeat) |
| Evans et al, 2010 (ON) | RV | DDS | 20 | 20 | | 12.7 | | | | | -1.13 | 0.12 | Card arranging reward responsivity objective test | Reward responsivity |
| Drew et al, 2020 (ON) | RV | ICD | 26 | 23 | | 2.78 | | | | | -0.48 | 0.08 | Rewarded speed of gaze shifting task | Residual velocity reward sensitivity |
| Lawrence et al, 2011 (ON) | RV | Apathy | 10 | 10 | | 0.38 | | | | | 0.20 | 0.28 | Rewarded spatial search task | Reward related speeding |

### Supplement table 6. Study summary statistics, tasks and measures included in the Meta-Analysis of Parkinson's on and off dopaminergic medication

| Reward processing summary statistics for Parkinson's disease patients ON vs OFF dopaminergic medication | | | | | | | Tasks and Measures Included in the Meta-Analysis. | |
|---|---|---|---|---|---|---|---|---|
| Author, Year | Category | N | t | F | d | Vard | Task | Measure |
| Czernecki et al, 2002 | RL | 22 | | 0.05 | 0.048 | 0.046 | Iowa Gambling Task | Number of advantageous minus disadvantageous choices |
| Graef et al, 2010 | RL | 14 | -0.374 | | -0.100 | 0.072 | Instrumental learning task | % correct choices with constant reward contingencies |
| Cavanagh et al, 2017 | RL | 28 | | 2.87 | 0.320 | 0.0375 | Value of volition task | % selection of A (90% reward) vs C (70% reward) – B (10% reward) vs D (30% reward) stimuli |
| Bodi et al, 2009 | RL | 26 | | 13.47 | 0.720 | 0.0385 | Feedback-based probabilistic classification task | Pattern of optimal decision selection over time |
| Chong et al, 2015 | OV | 26 | | 25.9 | 0.998 | 0.058 | Effort based decision making task (apple gathering) | Effort indifference point |
| Kobayashi et al, 2019 | OV | 24 | | 7.48 | 0.558 | 0.048 | Economic choice task | Relative risk aversion coefficient |
| Le Heron et al, 2018 | OV | 39 | 2.45 | | 0.392 | 0.028 | Effort based decision making task (apple gathering) | Mean difference in proportion of offers accepted |
| Mcguigan et al, 2018 | OV | 20 | 3.05 | | 0.682 | 0.062 | Cogntiive effort task | Mean difference in k-value |
| Cools et al, 2003 | OV | 12 | | 5.9 | 0.701 | 0.104 | Incentivised decision making task | Difference score of bets places in ascending and descending conditions. |
| Le Bouc et al, 2016 | OV | 20 | 2.76 | | 0.617 | 0.060 | Effort based decision making task: Binary choice task | Choice of effort level at the highest stake level. |
| Sharp et al, 2013 | OV | 18 | 1.17 | | 0.276 | 0.058 | Vancouver gambling task | Adjusted y intercept |
| Timmer et al, 2018 | RV | 23 | | 3.031 | 0.363 | 0.046 | Rewarded task switching paradigm | Reward related speeding 'repeat' condition |
| Muhammed et al, 2016 | RV | 30 | | 10.8 | 0.600 | 0.039 | Rewarded speed of gaze shifting task | Reward sensitivity in peak velocity |
| Evans et al, 2010 | RV | 38 | | 0.4 | 0.103 | 0.026 | Card arranging reward responsivity objective test | Reward responsivity |
| Drew, 2020 | RV | 26 | | 5.178 | 0.446 | 0.042 | Rewarded speed of gaze shifting task | Residual velocity reward sensitivity |

**Modified version of the Newcastle-Ottawa Scale for Assessing the Quality of Nonrandomized Studies in Meta-Analyses, used to assess potential sources of bias.**

Potential sources of bias were assessed using a modified version of the Newcastle-Ottawa Scale for Assessing the Quality of Nonrandomized Studies in Meta-Analyses. The studies are scored on:

**1. PD Definition: Is the case definition adequate?**

A) Cases were defined as PD according to a validated assessment tool/criteria or by an experienced clinician
B) Cases were defined as PD according to a validated assessment tool/criteria but the method for assessing PD status was not stated.
C) Cases were described as 'clinically' but no further description was given.

**2. PD Generality: Was a General sample of cases tested?**

A) A General sample of PD was tested.
B) Recruitment of PD cases was restricted to a specific sub-sample (specific age range, hospitalised only etc.)

**3. HC Selection: Selection of Controls**

A) Controls were selected from the same population as cases
B) Controls were not selected from the same population as cases
C) No description

**4. HC Definition: Definition of Controls**

A) HC were defined clearly defined as having no current or past psychopathology
B) Controls were not clearly defined as having no current or past psychopathology.

**Comparability (Comparability of cases and controls on the basis of the design or analysis)**

1. Does the study control for Age: Yes/No/Unclear
2. Does the study control for Gender: Yes/No/Unclear
3. Does the study control for IQ: Yes/No/Unclear
4. Does the study control for Socioeconomic status: Yes/No/Unclear
5. Does the study control for PD severity: Yes/No/Unclear
6. Does the study control for medication status: Yes/No/Unclear

Key: A, B, C
Y = yes, N= no, N/A= Not applicable (such as if no healthy control group in study)

### Supplement table 7. Quality rating of included studies

| Study quality ratings | | | Modified version of the Newcastle-Ottawa Scale | | | | | | | | | |
|---|---|---|---|---|---|---|---|---|---|---|---|---|
| Study | Year | Category | PD Definition | PD Generality | HC Selection | HC Definition | Age | Gender | IQ | Socioeconomic status | PD severity | PD medication |
| Bayard[1] | 2016 | OV | A | A | B | A | Y | Y | N | N | Y | Y |
| Brandt[2] | 2015 | OV | A | A | C | B | Y | Y | Y | N | Y | Y |
| Chong[3] | 2015 | OV | B | A | A | A | Y | Y | N | N | Y | Y |
| Cools[4] | 2003 | OV | A | A | C | B | Y | N | Y | N | Y | Y |
| Haagensen[5] | 2020 | OV | C | A | A | B | Y | Y | N | N | Y | Y |
| Kobayashi[6] | 2019 | OV | A | A | C | B | Y | Y | N | N | Y | Y |
| Le Bouc[7] | 2016 | OV | C | B | A | B | Y | Y | N | N | Y | Y |
| Le Heron[8] | 2018 | OV | A | A | A | A | Y | Y | N | N | Y | Y |
| Mcguigan[9] | 2018 | OV | A | A | A | A | Y | Y | N | N | Y | Y |
| Torta[10] | 2009 | OV | A | B | C | B | Y | Y | N | N | Y | Y |
| Sharp[11] | 2013 | OV | A | A | C | B | Y | N | N | N | Y | Y |
| Voon[12] | 2011 | OV | A | A | n/a | n/a | Y | Y | N | N | Y | Y |
| Balconi[13] | 2018 | RL | A | A | n/a | n/a | Y | N | N | N | Y | Y |
| Balusubramani[14] | 2015 | RL | C | A | C | A | N | N | N | N | Y | Y |
| Bentivoglio[15] | 2012 | RL | A | A | n/a | n/a | Y | Y | Y | N | Y | Y |
| Biars[16] | 2019 | RL | C | B | n/a | n/a | Y | Y | Y | N | N | Y |
| Bodi[17] | 2009 | RL | C | B | C | A | Y | N | Y | Y | Y | Y |
| Buelow[18] | 2014 | RL | A | A | A | A | Y | Y | Y | N | Y | Y |
| Castrioto[19] | 2015 | RL | C | B | C | B | Y | Y | N | N | Y | Y |
| Cavanagh[20] | 2017 | RL | C | A | C | B | Y | Y | Y | N | Y | Y |
| Czernecki[21] | 2002 | RL | A | B | B | A | Y | Y | N | N | Y | Y |
| Delazer[22] | 2009 | RL | A | A | C | A | Y | Y | N | N | Y | Y |
| Euteneuer[23] | 2009 | RL | C | A | C | B | Y | Y | N | N | Y | Y |
| Evens[24] | 2015 | RL | A | A | A | A | Y | Y | N | N | Y | Y |
| Garofalo[25] | 2017 | RL | A | A | C | B | Y | Y | Y | N | Y | Y |
| Gescheidt[26] | 2012 | RL | A | A | C | A | Y | Y | N | N | Y | Y |
| Graef[27] | 2010 | RL | A | B | B | A | Y | Y | N | N | Y | Y |
| Herzallah[28] | 2017 | RL & RV | B | A | A | A | Y | N | N | N | Y | N |
| Housden[29] | 2010 | RL | C | A | A | A | Y | Y | Y | N | Y | Y |
| Ibarretxe-Bilbao[30] | 2009 | RL | A | A | A | B | Y | Y | N | N | Y | Y |
| Kobayakawa[31] | 2008 | RL | C | A | A | A | Y | Y | N | N | Y | Y |
| Kobayakawa[32] | 2010 | RL | C | A | A | A | Y | Y | N | N | Y | Y |
| Mapelli[33] | 2014 | RL | A | A | A | A | Y | Y | N | N | Y | Y |
| Martinez-Horta[34] | 2013 | RL | A | A | n/a | n/a | Y | N | N | N | Y | Y |
| Mimura[35] | 2006 | RL | C | A | B | A | Y | Y | N | N | Y | Y |
| Pagonabarraga[36] | 2007 | RL | A | A | A | A | Y | Y | N | N | Y | Y |
| Paz-Alonso[37] | 2019 | RL | A | A | A | B | Y | Y | Y | N | Y | Y |
| Perretta[38] | 2005 | RL | A | A | A | A | Y | Y | N | N | Y | Y |
| Pignatti[39] | 2012 | RL | A | B | C | B | N | N | N | N | N | N |
| Pineau[40] | 2016 | RL | A | A | n/a | n/a | Y | Y | N | N | Y | Y |
| Piray[41] | 2014 | RL | C | B | C | A | Y | N | Y | N | Y | N |
| Poletti[42] | 2010 | RL | A | A | C | B | Y | Y | N | N | Y | Y |
| Poletti[43] | 2011 | RL | A | B | n/a | n/a | Y | Y | N | N | N | Y |
| Rossi[44] | 2010 | RL | A | A | n/a | n/a | Y | Y | N | N | Y | Y |
| Sáez-Francàs[45] | 2014 | RL | C | A | n/a | n/a | Y | Y | N | N | Y | N |
| Thiel[46] | 2003 | RL | C | A | C | A | Y | N | N | N | Y | Y |
| Timmer[47] | 2017 | RL | A | A | A | A | Y | Y | Y | N | Y | Y |
| Xi[48] | 2015 | RL | A | B | C | B | Y | Y | N | N | Y | Y |

| | | | | | | | | | | | | |
|---|---|---|---|---|---|---|---|---|---|---|---|---|
| Yildirim[49] | 2020 | RL | A | A | B | A | Y | Y | N | N | Y | Y |
| Drew[50] | 2020 | RV | A | B | C | A | Y | Y | N | N | Y | Y |
| Evans[51] | 2010 | RV | A | A | B | B | Y | Y | N | N | Y | Y |
| Lawrence[52] | 2011 | RV | A | A | n/a | n/a | Y | N | N | N | Y | Y |
| Muhammed[53] | 2016 | RV | C | A | B | B | Y | Y | Y | N | Y | N |
| Renfroe[54] | 2016 | RV | B | B | A | A | Y | Y | N | N | Y | Y |
| Timmer[55] | 2018 | RV | A | A | A | B | Y | N | Y | N | Y | Y |
